# Resistance exercise training for skeletal muscle health in adults living with multiple long-term conditions: a scoping review

**DOI:** 10.1101/2025.09.17.25335988

**Authors:** Christopher Hurst, Jonas Johansson, Miles D Witham, Sian M Robinson, Rachel Cooper, Avan A Sayer

## Abstract

Low muscle strength is prevalent in individuals living with multiple long-term conditions (MLTC). Resistance exercise (RE) training is the most effective treatment for improving strength and physical performance across the lifecourse, but little is known about RE in people living with MLTC. This scoping review aimed to identify published evidence on RE as a treatment for improving muscle mass, strength and physical performance in adults living with MLTC. Six electronic databases were searched to identify studies that included adults aged >18 years who were living with two or more long-term conditions and undertaking a RE intervention. Sixty-eight studies met the inclusion criteria. A range of methods for characterising MLTC were reported; almost half of the studies (30/68) that included people living with ≥2 LTC focused on co-morbidity (i.e., participants selected based on the presence of an index condition plus one or more co-existing conditions). Only seven studies specifically described their population as living with MLTC. Most studies involved RE within a multicomponent intervention with considerable variability in exercise programming. Of the studies which assessed lean mass or muscle mass (n=17), only one reported a post-intervention increase. Muscle strength improved in 47% (16/34) of studies that examined this, with improvements in physical performance reported across most studies (82%). Exercise session attendance was generally good with few reported adverse events. RE training may be effective for improving strength and physical performance in adults living with MLTC. Importantly, RE was well tolerated with good adherence and low numbers of adverse events reported.

## 1 Introduction

Multiple long-term conditions (MLTC), also known as multimorbidity, typically refers to the co-existence of two or more physical or mental health conditions lasting 12 months or more [1]. The number of individuals living with MLTC has increased substantially in recent decades and it is now recognised as one of the biggest challenges facing modern medicine [2–4]. People living with MLTC are the main recipients of healthcare and as MLTC increases in prevalence with age it is the norm for those aged 65 and above [2,5]. The presence of MLTC is associated with a range of adverse outcomes including reduced quality of life, loss of physical independence, higher rates of emergency hospital admission and increased rates of all-cause mortality [6–9].

Low muscle strength influences an individual’s ability to remain physically independent and is prevalent in individuals living with MLTC even after taking account of age [10]. In addition, those living with MLTC are more likely to experience accelerated loss of muscle strength over time compared with people without MLTC [11], especially if they are living with certain combinations of conditions [12]. Low muscle strength currently represents the primary consideration in the diagnosis of sarcopenia [13], a progressive and generalised skeletal muscle disorder that involves the loss of muscle mass and strength with age [14,15]. The presence of sarcopenia results in impaired physical functioning, loss of physical independence and reduced quality of life [14,16]. Given the high prevalence of sarcopenia in people living with MLTC [12], attention needs to be paid to developing strategies to prevent and treat sarcopenia in this growing segment of the population.

While previous research has suggested that exercise training is safe and has the potential to be beneficial for those living with MLTC [17,18], not all exercise is equally effective for maintaining and improving muscle strength and physical performance [19]. Resistance exercise (RE) training is the most effective strategy for improving skeletal muscle mass and strength across the life course [20,21] and is currently recommended as the first-line treatment for sarcopenia in clinical practice [15,22,23]. Moreover, the benefits of RE across a range of conditions including cardiovascular diseases [24], pulmonary diseases and musculoskeletal disorders are well documented [25] and highlight the potential of this exercise mode as a treatment strategy. Despite this, the specific effects of RE on muscle size, strength and physical performance in adults living with MLTC remain largely unknown. This knowledge gap is compounded by the fact that participants living with MLTC are often excluded from trials including those of exercise, which often focus on younger adults among whom MLTC would not be expected to be so prevalent. Understanding these potential effects, as well as which exercise programme characteristics are associated with improved outcomes, will support the design and delivery of effective exercise programmes that meet the needs of adults living with MLTC [26,27].

The combined illness and treatment burden experienced by those living with MLTC, who may interact with numerous clinical services, can add substantial complexity to the prescription and delivery of exercise [26,28]. As such, understanding more about the feasibility and potential effectiveness of RE is an important first step to support the design and delivery of evidence-based exercise programmes tailored to the needs of those living with MLTC [29]. Therefore, the aim of this scoping review was to identify and summarise published evidence evaluating RE training as a strategy for maintaining and/or improving skeletal muscle mass, strength and physical performance in adults living with MLTC.

## 2 Methods

We conducted a scoping review according to a prespecified protocol, informed by the framework for scoping reviews described by Arksey and O’Malley [30]. This is reported in accordance with the Preferred Reporting Items for Systematic Reviews and Meta-analyses extension for Scoping Reviews (PRISMA-ScR) [31]. The completed PRISMA-ScR checklist is available in Table S5. The protocol for this scoping review has not previously been published.

### 2.1 Eligibility criteria

Eligibility criteria were formulated using the Participants, Intervention, Comparator, Outcomes and Study Design (PICOS) framework. Studies were considered eligible for inclusion if they met the criteria outlined in Table 1 and were peer-reviewed original research articles published in the English Language.

**Table 1.** Inclusion and exclusion criteria.

| Category | Inclusion criteria | Exclusion criteria |
| --- | --- | --- |
| Participants | Adults aged $\geq 18$ years who were living with two or more long-term conditions. | Studies which included only a Charlson comorbidity score were excluded as the score alone does not allow analyses of whether multiple conditions are present. |
| Intervention | Studies which included a resistance exercise (RE) intervention $\geq 2$ weeks in duration consisting of either:<br>1. RE training alone<br>2. RE completed as part of a multi-component exercise intervention (i.e., combined with other exercise modes)<br>3. RE completed as part of a wider multi-component intervention (e.g., combined with dietary or other interventions).<br>Studies delivering RE in person, virtually, or via hybrid methods. | Studies which provided only guidance on RE training but did not include at least one contact with a trained professional during the intervention period. |
| Comparator | Studies with or without a control or comparator group. Single group studies were required to have measured outcomes at baseline and follow-up (i.e., pre-test, post-test) to be eligible for inclusion. |  |
| Outcomes | Assessment of at least one of the following outcomes based on the revised European Working Group on Sarcopenia in Older People (EWGSOP2) definition for sarcopenia [13]<br>1. Muscle mass or size measurement (e.g., MRI, CT); lean mass measurement (e.g., DXA)<br>2. Muscle strength assessment (e.g., 1-RM, handgrip strength)<br>3. Physical performance (e.g., 5-repetition chair stand, gait speed, SPPB, TUG) | Studies which provided only results on feasibility, tolerance, and/or adherence. |
| Study | Experimental or quasi-experimental | Observational studies, retrospective |
| design | randomised (e.g., RCTs, or pilot RCTs) or non-randomised trials and single group, pre-test, post-test studies. | analyses of patient databases, service audits or secondary analyses of registry data were not included. |
| *CT* Computed Tomography, *DXA* Dual-energy X-ray absorptiometry, *MRI* Magnetic Resonance Imaging, *RCT* Randomised Controlled Trial, *SPPB* Short Physical Performance Battery, *TUG* Timed up-and-go, *1-RM* 1-Repetition Maximum |  |  |

Due to the relatively recent operationalisation of MLTC, combined with the ongoing variation and lack of consistency in defining MLTC [32] we considered MLTC to be present when: 1) authors identified the study participants as living with MLTC or 2) participants were living with two or more long-term conditions, as per widely used definitions of MLTC [1], or participants were living with an index condition plus ≥50% of the study sample were living with at least one other long-term condition (more often referred to as co-morbidity) or 3) over 50% of the participants were living with two or more long-term conditions as identified in the baseline characteristics of the study sample.

### 2.2 Information sources and search strategy

The following electronic databases were searched from inception until 1^st^ May 2025: MEDLINE (via Ovid), Embase (via Ovid), Cumulative Index to Nursing and Allied Health Literature (CINAHL), The Cochrane Central Register of Controlled Trials (CENTRAL), Scopus (Elsevier Science Publishers) and Web of Science (Institute for Scientific Information). A combination of free text and MeSH terms were used which were initially applied in the MEDLINE database and then modified for use according to the specifications of each database (see Table S1 for the complete search strategy). We also searched two clinical trial registries 1) ISRCTN (https://www.isrctn.com) and 2) ClinicalTrials.gov (https://clinicaltrials.gov) to identify potentially relevant studies and any associated publications. We examined reference lists of included studies and performed backward citation tracking to identify other potential studies for inclusion.

### 2.3 Study selection

All retrieved search results were exported to a specialist citation management software programme (Zotero 6.0) before being transferred to Rayyan (www.rayyan.ai). Potential duplicates were identified using the automated duplicate detection tool in Rayyan before being checked manually by the lead author. All titles and abstracts were then independently screened by two reviewers (CH, JJ). Following this, the full texts of all potentially eligible records were retrieved and screened against our predetermined inclusion and exclusion criteria by the same reviewers to identify those papers eligible for inclusion in the review. Any disagreements between reviewers during the title/abstract and full-text screening stages were resolved by discussion with the wider author group.

### 2.4 Data extraction

Data were extracted from included studies using a standardised proforma. Two investigators (CH & JJ) independently extracted data from all studies which were then checked for accuracy and completeness. Extracted data included (1) study characteristics, (2) participant characteristics, (3) RE intervention characteristics and (4) study outcomes. A formal quality assessment was not performed as this is outside the remit of a scoping review [30].

### 2.5 Data synthesis

Data are presented in tables and narratively summarised in the text. We did not perform a quantitative synthesis of the data because of the breadth of our review and the expected heterogeneity in reporting of study findings.

## 3 Results

### 3.1 Search results

Database and registry searching yielded a total of 5186 unique records. Following title and abstract screening, 556 records were retrieved for full-text review and of these 61 were identified as eligible for inclusion. Registry searching identified 38 records, but screening of these records did not identify any further eligible studies. A further eight studies potentially eligible for inclusion were identified through searching of reference lists of included studies and 7 of these were classified as eligible for inclusion after full-text review. In total, 68 studies were included in this review [33–100]. A flow diagram for study identification is shown in Figure 1.

**Figure 1.**
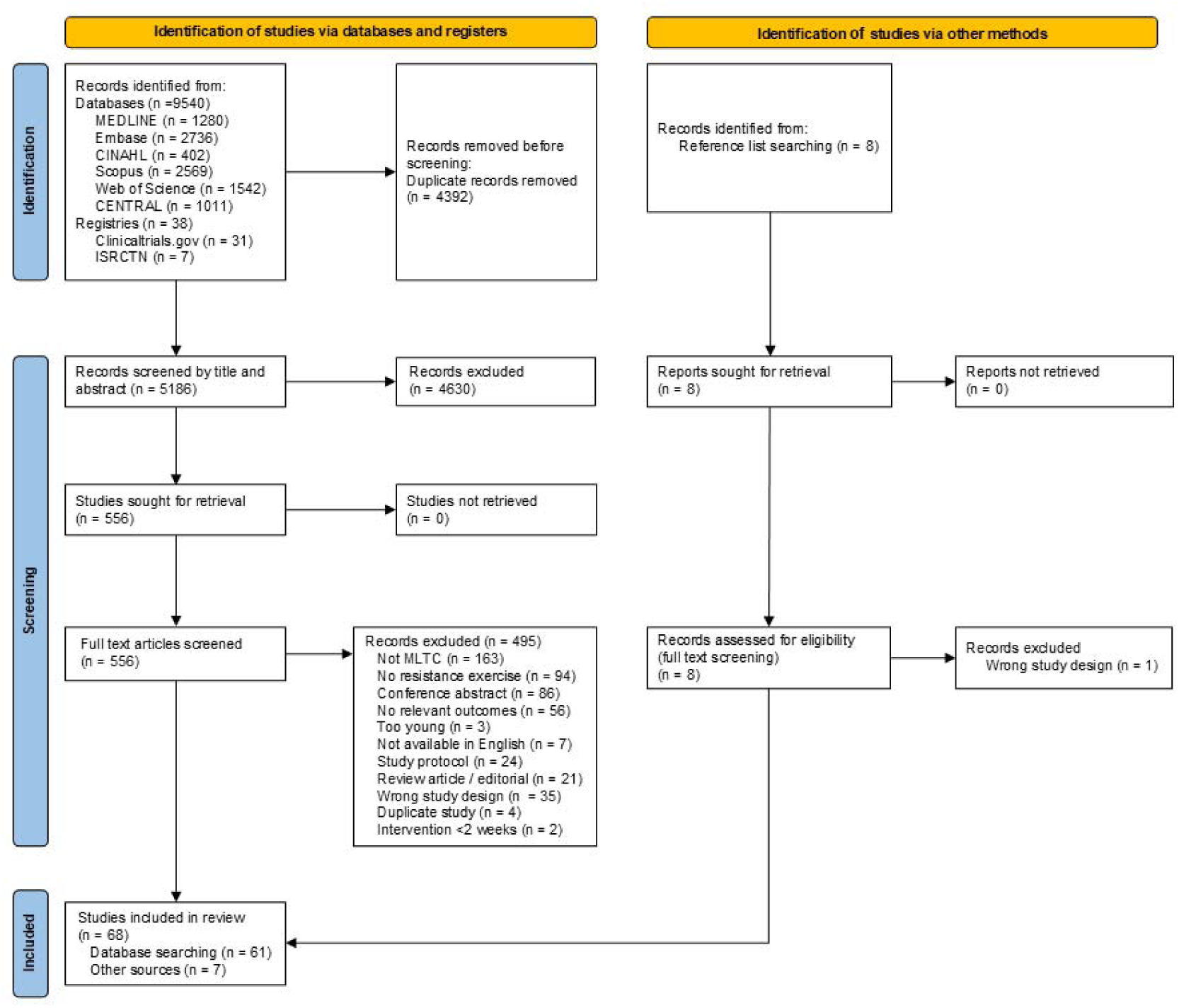
PRISMA flow chart.

### 3.2 Study characteristics

Included studies consisted of RCTs (n = 42), non-randomised controlled trials and parallel groups trials (n = 14), as well as single group, pre-post studies (n = 12). Studies were conducted in different world regions including Europe, North America, South America, Asia, and Oceania with the highest number of included studies conducted in the USA (n = 22).

Sample sizes ranged from 4 to 964 participants although 37% of studies included <50 participants and only two studies involved >500 participants. Most of the included studies (58/68) involved both male and female participants. The mean age of participants in the included studies ranged from 29 to 82 years with most studies (∼75%) having a mean age of over 65 years. Studies were published from 1994 to 2025 with the majority (79%) published in the last 10 years; 53% of studies were published from 2020 onwards. A more detailed summary of the characteristics of the included studies can be found in Table S2.

### 3.3 Approaches to defining and identifying MLTC

Methods used for identifying MLTC within the included studies are summarised in Table 2. The most common method (30/68 studies) for identifying those living with ≥2 LTC focused on co-morbidity (i.e., participants selected based on the presence of an index condition plus one or more co-existing condition). A wide range of index conditions were studied with the most common being: cancer (n = 7), CKD (n = 5), osteoarthritis (n = 4), CHF (n = 4) and COPD (n = 4). Only seven studies (∼10%) were described specifically by the authors as including adults living with MLTC and 23 studies (34%) were included based on a count of conditions being ≥2.

**Table 2.** Summary of methods used for identifying MLTC.

| Method used | Number of studies | References |
| --- | --- | --- |
| Author defined MLTC | 7 | [68,78,80,83,85,89,95] |
| ≥2 conditions based on count | 23 | [34,35,39,41,46,53,55,57–59,61–66,72–74,77,86,87,96] |
| Index condition + additional conditions |  |  |
| Cancer | 7 | [51,54,60,69,79,90,98] |
| CHF | 4 | [49,52,81,88] |
| CKD | 5 | [36,56,71,92,97] |
| COPD | 4 | [38,40,44,50] |
| Diabetes | 2 | [42,84] |
| HIV | 1 | [75] |
| Osteoarthritis | 4 | [43,67,70,76] |
| Peripheral arterial disease | 1 | [47] |
| Rheumatoid arthritis | 1 | [93] |
| Stroke | 1 | [82] |
| Specific combination of conditions |  |  |
| COPD & CHF | 1 | [37] |
| Diabetes and Osteoarthritis | 2 | [33,45] |
| Scores and Indices |  |  |
| Davies Comorbidity Score | 1 | [100] |
| Functional Comorbidity Index | 4 | [48,91,94,99] |
| *CHF* Chronic Heart Failure, *CKD* Chronic kidney disease, *COPD* Chronic Obstructive Pulmonary Disease, *HIV* Human immunodeficiency virus, *MLTC* Multiple long-term conditions |  |  |

Most studies did not indicate which conditions participants were asked to consider when self-reporting the presence of long-term conditions. There were exceptions to this however, with Barker et al. [34] using a list of 40 long-term conditions. Further detail on the approaches used to characterise MLTC within the included studies can be found in Table S2.

### 3.4 Resistance exercise programme design

There was considerable variation in the design and delivery of RE programmes (see Table S3 for details). Of the included studies, only 12 (18%) included RE alone, with most involving RE performed as part of a multicomponent exercise programme. Aerobic exercise was included in ∼70% of studies, with balance, mobility, and flexibility training also frequently performed. Several studies involved a multicomponent intervention combining other elements with exercise, including educational activities [76,85], cognitive training [68], a weight management programme [42], smoking cessation therapy [47] or dietary/nutritional manipulation [71,96].

Exercise programmes ranged from 4 weeks to 24 months in duration, although two prehabilitation studies included interventions where some participants were engaged for less than 4 weeks due to time limitations prior to surgery [69,79]. Most studies involved individual RE training sessions that lasted 30-60 minutes and were typically performed 1-3 times per week, although some studies involved >3 sessions per week [79,87,89]. In several studies, participants were asked to perform home-based exercise on non-training days. A wide range of RE intensities were prescribed across the included studies with methods including one-repetition maximum (1RM) and Rating of Perceived Exhaustion (RPE) used. However, the RE intensity was often imprecisely described, difficult to interpret, or not reported at all. Most studies reported that the training stimulus provided to participants was progressed over the duration of the intervention by increasing volume or intensity, by increasing load or changing the resistance band that was used. However, several studies did not report how the RE training was progressed across the intervention period and it was unclear if any progression occurred at all.

RE was performed using free-weights, machines and body weight exercises, and was delivered across a broad range of settings including in hospital or other clinical environments (e.g., outpatient clinic), community facilities and at the participants’ homes. Several studies combined delivery settings, for example beginning the intervention within a hospital/clinic before progression to unsupervised, home-based exercise. Exercise was delivered both individually and in groups and was often supervised by physiotherapists or other qualified exercise practitioners. Most studies made no mention of whether RE was tailored or adapted for specific LTC, and if so how this was done, although there were exceptions. For example, in the study from Barker et al. [34], clinicians were encouraged to individualise the exercise programme to accommodate participants’ LTC, with 50% of participants requiring individual adjustments to their exercise program to accommodate their MLTC.

### 3.5 Outcome measures

Details on the outcome measures assessed within each included study can be found in Table S4. Quantification of muscle or lean mass was included in 17 of the included studies. Of these 17 studies, seven had used Dual-energy X-ray absorptiometry (DXA) [42,48,51,60,96,98,100], six had used bioelectrical impedance analysis (BIA) [46,50,53,54,66,90], two had used computed tomography [60,71], one study had used Magnetic Resonance Imaging [61] and one air displacement plethysmography [62], and a final study did not report the technique used [76].

Thirty-four (50%) of the included studies measured muscle strength. Grip strength was the most commonly used method (n = 27); 1RM (n = 7), 10RM (n = 2) and isometric strength assessment of the lower body (n = 5) were also reported. Several studies [52,56,73,76,82,83,89,91] assessed muscle strength using more than one method (e.g., grip strength and 1RM) and nine studies assessed muscle strength, typically 1RM or 10RM, across more than one exercise or muscle group (e.g., leg press and chest press).

Physical performance was measured in 81% (55/68) of the included studies with 34 studies including more than one method of assessment. A range of assessments were used to evaluate physical performance including: 6-minute walk test (6MWT) (n = 30); Timed up-and-go (TUG) (n =26); gait speed (n = 18); Short Physical Performance Battery (SPPB) (n = 16); 30s chair stand test (n = 13); 5-repetition chair stand test (n = 10); 10 repetition chair stand (n = 1); 10m walk (n = 2) and 400m walk (n = 1).

There were 25 studies (37%) which included measurements of both strength and physical performance, and 10 studies (15%) which assessed muscle size, muscle strength and physical performance.

### 3.6 Effects of RE on muscle mass, strength, and physical performance

A summary of the main findings from the included studies is presented in Table 3 with further detail in Table S4.

**Table 3.**
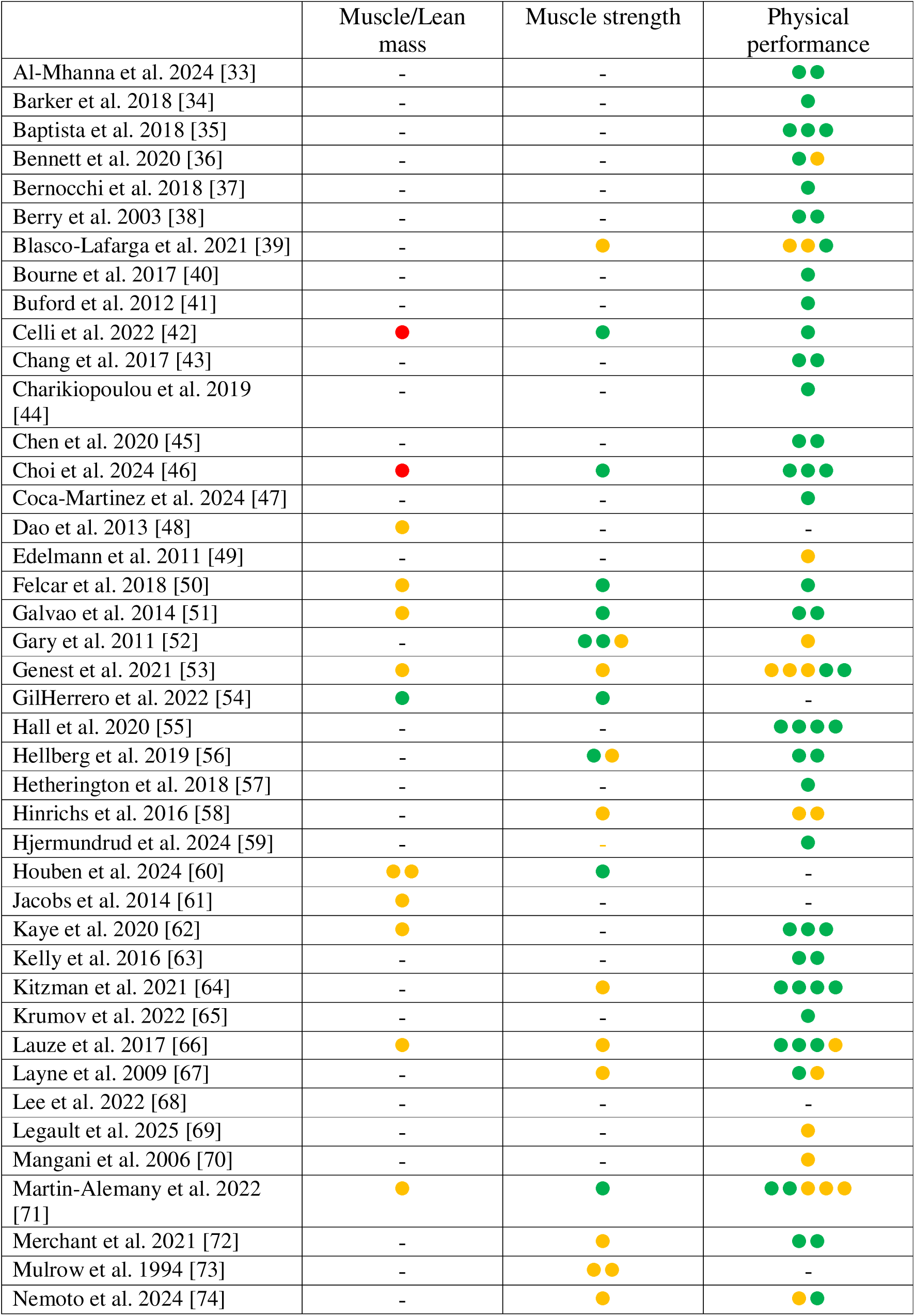
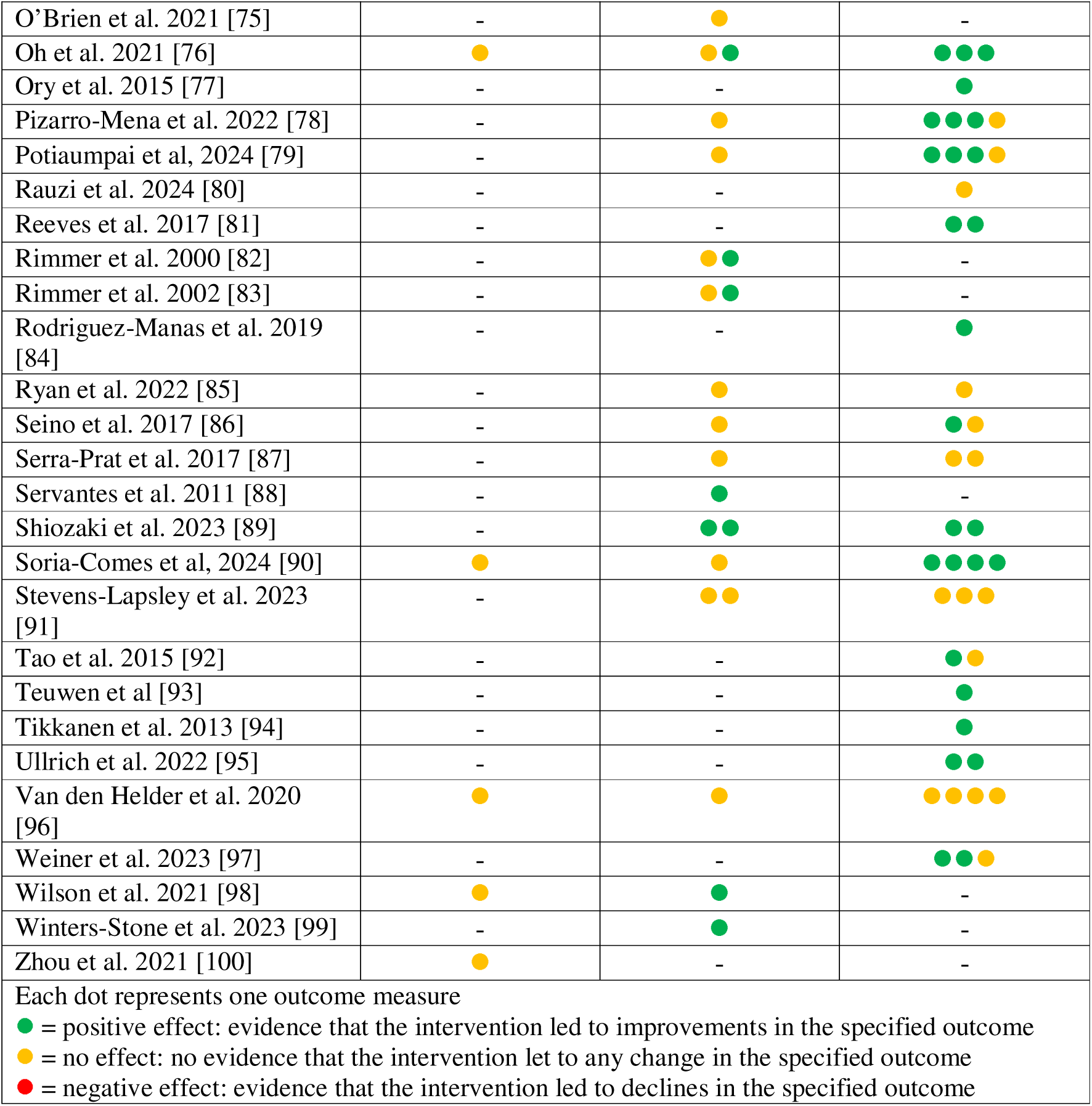
Summary of main findings from included studies (ordered alphabetically by first author surname).

Only one of the included studies reported an increase in lean or muscle mass [54]. In this study, GilHerrero and colleagues reported statistically significant increases in skeletal muscle mass estimated via BIA following a 12-week, twice-weekly multicomponent exercise programme (RE and aerobic exercise) in 14 people undergoing treatment for cancer. In contrast, the studies by Celli and colleagues [42] and Choi et al [46] reported a reduction in lean body mass assessed by DXA, and skeletal muscle mass assessed by BIA, respectively. In the study from Celli et al [42], the reduction in lean mass was accompanied by statistically significant weight loss, following a combined exercise and weight-management intervention. Although, Choi et al. reported a decline in muscle mass, the intervention induced change was small (<0.25kg) and a limited sample size (n = 7) precluded any statistical testing [46]. None of the other studies (14/17) that assessed muscle mass reported any post-intervention change.

Of the 34 studies that assessed muscle strength, 16 (47%) reported a beneficial effect. Increases in 1RM strength were reported consistently, but this method of assessment was only included in a small number of studies (n = 7). There were statistically significant improvements in 1RM strength assessed by a single movement [99] or across multiple movements [42,50,51,54,60,98]. Increases in muscle strength were also reported consistently across several other strength measures including 10RM [82,83], isometric lower-body strength [52,56,89] and isokinetic strength [88]; although improvements in strength using these methods were not observed in every study [73]. Only three of the studies which assessed grip strength (n = 27) reported statistically significant post-intervention improvements [52,71,89]. Two of the included studies demonstrated improved upper- and lower-body strength although simultaneously reported no change in grip strength [82,83].

Most of the studies (82%) which assessed physical performance reported improvements across a range of outcome measures. However, improvements in physical performance were not universal with a number of studies (n = 12) reporting improvements in some measures of physical performance while simultaneously reporting no change in others. Ten of the included studies reported no improvements in physical performance. There was considerable variation in these studies with no effect reported across several outcomes including 6MWT [49,52,70], TUG [58,87] and SPPB [91,96]. Seven studies which reported improvements in physical performance also reported concomitant improvements in muscle strength [42,50,51,56,71,76,89]. There were 5 studies where there was no improvement in either physical performance or strength [58,85,87,91,96].

### 3.7 Adverse events, adherence, and intervention fidelity

Information relating to adverse events was reported in 45 (66%) of the 68 included studies. Of those studies which provided details, 22 (49%) reported no adverse events related to the exercise intervention. Several studies reported the occurrence of minor adverse events including musculoskeletal pain exacerbations, exercise induced muscle soreness, chest pain, dizziness, and falls. A small number of included studies noted the occurrence of serious adverse events which represented 14-40% of the reported adverse events [58,81,91,96,97]. These included one myocardial infarction [81], although the majority were not described. Nearly all of these were described as unrelated to the exercise intervention and were primarily exacerbations of pre-existing conditions.

There was limited detail provided about adherence and intervention fidelity, although most studies did report some information relating to exercise session attendance. Adherence to an intervention was typically reported in terms of the proportion of exercise sessions that were attended by the participants, primarily focusing on supervised exercise sessions. Some authors used qualitative descriptors of adherence such as ‘low’ [76] or adhering ‘well’ to an exercise intervention [87]. There was considerable variation in exercise session attendance across the included studies, with attendance ranging from below 70% to greater than 90% attendance. In the studies that provided information, there was substantial variation in attendance between participants within a study. Further details are reported in Table S4.

## 4 Discussion

This scoping review has identified and summarised published evidence evaluating RE as a strategy for improving skeletal muscle mass, strength and/or physical performance in adults living with two or more LTC. The 68 studies identified report on a range of diverse RE programmes delivered across different settings. Most of these programmes combined RE with other exercise modes or with other types of intervention. Only one study reported an improvement in lean mass, but improvements in muscle strength, typically lower-body strength, and physical performance were more commonly observed although not consistently; muscle strength improved in around half of the studies where it was measured with improvements in physical performance reported in 82%. More studies which have assessed all 3 outcomes (muscle mass, muscle strength and physical performance) are needed to determine if the effects of RE vary across these outcomes. Importantly, only a relatively small number of severe adverse events occurred, typically outside of the exercise sessions, with participants willing and able to engage in the exercise. This suggests that RE training is safe and tolerable in populations living with MLTC if programmed and implemented appropriately.

This synthesis of studies investigating the potential of RE in adults living with MLTC represents an important step forward in this area and highlights considerable progress in the field since a previous review in 2012 which suggested that those living with MLTC can improve mobility and physical functioning via exercise [101]. Since then, the role of exercise for adults living with MLTC has received increasing attention with evidence demonstrating that exercise is feasible and effective in this group [17,102]. Despite this progress, there has been only limited consideration of the effect of exercise on skeletal muscle-related outcomes in the context of MLTC. Sarcopenia is commonly observed in adults living with MLTC highlighting the importance of identifying and evaluating potential interventions in this population [10]. The diversity of MLTC patterns we have observed within our included studies (e.g., cancer patients, heart failure patients, community dwelling older adults) makes it challenging to fully interpret the effects of RE on skeletal muscle-related outcomes. Living with MLTC can make it harder to engage in exercise due to symptoms including pain, breathlessness and dizziness with different patterns of MLTC likely resulting in different responses to exercise [89]. Variation in treatment approaches and the effect of treatments, including drug induced side-effects, across different long-term conditions also need to be considered. These factors highlight the need for more nuanced exercise trials involving those living with MLTC which could help to identify appropriate strategies for adapting exercise to meet the individual requirements resulting from the presence of specific combinations of LTC.

The findings of our review suggest that RE has potential to improve muscle strength and physical performance in adults living with MLTC. Previous work has shown that RE is the most effective exercise mode for improving muscle strength and physical function with beneficial effects consistently reported across a range of LTC [103,104]. Although our findings suggest potential beneficial effects of RE, considerable variation in participant characteristics (including the diversity of LTC included) and exercise programme characteristics mean that it is important not to overinterpret our findings. Further data, drawn from well powered controlled trials is needed before making definitive statements about the efficacy of RE across muscle-related outcomes. Many of the included studies were small and so limited statistical power may have contributed to the finding of non-significant results in a number of studies. This is likely to be of particular relevance in MLTC studies where the population is heterogeneous in terms of conditions and possibly function, whereby there is likely to be more random error and higher dropout (due to concomitant illness). These factors all mean that larger sample sizes are needed to reliably detect treatment effects. Future work is needed to evaluate these effects and understand how responses to exercise programmes vary depending on the presence (or absence) of specific LTC. This could include focusing on the evaluation of responses to minimal dose RE [105] which may be appealing in populations living with MLTC because of their existing treatment and condition burdens.

Most studies included in this review prescribed RE as part of a multicomponent intervention. This makes distinguishing the effects of RE alone on muscle strength and physical performance challenging as other exercise modes, including aerobic exercise, can have beneficial effects on skeletal muscle [106,107]. It is important to note however, that our scoping review did not set out to interrogate the size of effects reported, nor to compare effects between studies and therefore there remains a need to provide guidance around the content of effective RE programmes to adults living with MLTC which is often sub-optimally delivered in clinical practice [108]. Adults living with MLTC represent a broad group of people who experience a wide range of individual LTC which could achieve meaningful benefit from different modes of exercise (e.g., aerobic exercise for Heart Failure or COPD). The multi-system benefits of exercise across multiple tissues and organ systems could help to avoid polypharmacy and the associated potentially harmful drug interactions in those living with MLTC, thereby reinforcing the appeal of exercise as a treatment strategy in this population.

Importantly, the studies included in this review suggest that RE is a safe and viable exercise strategy for adults living with MLTC. This was illustrated by good attendance in most studies, although there was some variability between and within studies, and a low number of reported adverse events. Our findings are consistent with those from a previous systematic review by Bricca and colleagues who found that exercise therapy was not associated with an increased risk of adverse events [17]. Drawing on findings from a qualitative interview study, those living with MLTC are willing to engage with RE if they are appropriately supported to do so [26]. This reinforces the need for researchers, and those delivering exercise in clinical practice and beyond, to focus on the factors, including behaviour change techniques [109], to support engagement and adherence in this population.

### Gaps in the knowledge base and future considerations

Despite the positive findings presented in this review, further work is needed to fully elucidate the potential of RE for adults living with MLTC and to translate this knowledge into the delivery of effective exercise programmes in clinical practice and beyond.

Incomplete reporting of exercise interventions in some of our included studies makes it hard to draw inference about which RE characteristics are most consistently associated with improved outcomes. We would encourage authors to ensure clear and complete reporting in relation to exercise prescription so that research findings can be interpreted fully by the reader, and/or reproduced by the scientific community. The Consensus on Exercise Reporting Template (CERT) provides a useful guide here [110]. Authors should also strive to provide an assessment of intervention fidelity (i.e., did the participant complete the exercise as intended), to enable a more thorough evaluation of the intervention [111].

We would encourage authors of future studies to carefully consider their choice of outcome measures in the context of skeletal muscle strength and physical function, to pinpoint the effects of RE and the implications for populations living with MLTC. Outcome measures need to be selected which are responsive to the intervention under test. For example, despite the usefulness of grip strength as an indicator of overall health status and its relationship with a range of health outcomes [112], only three of 27 studies (11%) measuring grip strength in this scoping review showed beneficial effects. It may be that grip strength is not the most appropriate measure to evaluate changes in muscle strength following an exercise intervention (lower limb based RE is unlikely to meaningfully improve grip strength for example) [113]. There is also a need for an agreed set of core outcome measures for exercise studies in the context of MLTC. This should be informed by consideration of which domains are important to people living with MLTC, and how these domains should be measured. This is important in ensuring that a) the most informative evidence is collected, and b) outcomes can be compared or combined across studies.

More work is needed to elucidate the influence of RE in the context of specific LTC in those living with MLTC. For example, the consequences of treatment and how specific medications, conditions, or disease specific considerations—which vary substantially within the umbrella of MLTC— (e.g., consideration of the impact on painful joints in those with osteoarthritis or breathlessness in those with COPD) might contraindicate RE prescription as well as the physiological adaptations derived from exercise. There is a need to specifically address the management of these symptoms in the design of RE programmes to prevent these being a barrier to exercise engagement [114]. Further understanding could support the move towards a greater awareness of both the optimal and minimal dose of RE to improve skeletal muscle related outcomes in those living with MLTC. Issues relating to how RE is delivered (e.g. group-based or individual, supervised, or home-based), where it is delivered (e.g. gym, hospital, primary care, own home, community venues) and who it is delivered by (e.g. physiotherapists, specialist exercise practitioners, exercise physiologists, others) represent important considerations for future research. This future work should involve and engage patients and members of the public at every stage of the research process if benefits to people living with MLTC are to be realised.

Despite increasing attention, the measurement of MLTC is often poorly reported and highly variable in research [32], highlighting a need to improve consistency and transparency [115]. Different approaches to identifying and defining MLTC can make it challenging to synthesise findings from studies which include diverse populations. In addition to the challenges of defining MLTC, the included studies within this review have tended to focus on older adults but those at younger age living with MLTC are also affected by loss of muscle strength [10].

### Limitations

Despite a rigorous methodological process, this scoping review is not without limitations. Studies were included in the present review based on the criteria of involving participants with two or more LTC. This broad definition represents diverse patterns of LTC across various clinical contexts making it challenging to evaluate effects across studies within this population. It is possible that several potentially relevant studies were excluded as it was not possible to confirm the presence of two or more LTC based on the information provided by the authors. However, by keeping our inclusion criteria broad, it is possible that we may have included studies where the complete sample were not living with two or more LTC (i.e., we required >50% of participants to be living with ≥2 long-term conditions). Our interpretation of findings was largely based on statistical significance, with few studies evaluating findings in relation to clinical significance or smallest meaningful change.

## 5 Conclusion

This review identified 68 studies investigating RE in adults living with two or more long-term conditions. The findings suggest that RE may be effective for improving muscle strength and physical performance in adults living with MLTC. Importantly, RE appears to be well tolerated with good adherence and low numbers of adverse events reported suggesting that it may be a viable strategy for maintaining skeletal muscle health in adults living with MLTC. However, there is a need to improve both the identification of MLTC in exercise training studies and the reporting of exercise programmes to provide meaningful data that can be used to support the design and delivery of RE training as a treatment in clinical practice and beyond.

## Author Contributions

AAS conceived the idea for this work. All authors contributed to the design of the review. CH performed the literature search. CH and JJ screened articles for inclusion and extracted the data from relevant studies. CH and JJ drafted the article, which was critically revised by all other authors. All authors have read and approved the final version of this manuscript.

## Data Availability

All data produced in the present work are contained in the manuscript

## Acknowledgements

This work was supported by the National Institute for Health and Care Research (NIHR) Newcastle Biomedical Research Centre (reference: NIHR203309). The authors also acknowledge support from the ADMISSION research collaborative which is funded by the Strategic Priority Fund “Tackling multimorbidity at scale” programme [grant number MR/V033654/1]. This funding is delivered by the Medical Research Council and the National Institute for Health and Care Research in partnership with the Economic and Social Research Council and in collaboration with the Engineering and Physical Sciences Research Council. MDW acknowledges support from the NIHR Newcastle Clinical Research Facility. AAS, MDW and RC acknowledge support from the Multiple Long-term Conditions cross-NIHR collaboration.

The views expressed in this publication are those of the authors and do not necessarily reflect the views of the National Institute for Health and Care Research, the Department of Health and Social Care or UK Research and Innovation.

## Ethics statement

The authors have nothing to report.

## Consent

The authors have nothing to report.

## Supporting information

Additional supporting information can be found online in the Supporting Information section. Table S1: Full search strategy. Table S2: Characteristics of included studies. Table S3: Exercise intervention details. Table S4: Study outcomes. Table S5: Preferred Reporting Items for Systematic reviews and Meta-Analyses extension for Scoping Reviews (PRISMA-ScR) Checklist.

## References

1. Academy of Medical Sciences. Multimorbidity: A Priority for Global Health Research. Academy of Medical Sciences; 2018.

2. Barnett K, Mercer SW, Norbury M, Watt G, Wyke S, Guthrie B. Epidemiology of multimorbidity and implications for health care, research, and medical education: a cross-sectional study. The Lancet. 2012;380(9836):37-43. doi:10.1016/S0140-6736(12)60240-2

3. Whitty CJM, MacEwen C, Goddard A, et al. Rising to the challenge of multimorbidity. BMJ. 2020;368:l6964. doi:10.1136/bmj.l6964

4. Skou ST, Mair FS, Fortin M, et al. Multimorbidity. Nat Rev Dis Primers. 2022;8(1):1–22. doi:10.1038/s41572-022-00376-4

5. Cassell A, Edwards D, Harshfield A, et al. The epidemiology of multimorbidity in primary care: a retrospective cohort study. Br J Gen Pract. 2018;68(669):e245–e251. doi:10.3399/bjgp18X695465

6. Nunes BP, Flores TR, Mielke GI, Thumé E, Facchini LA. Multimorbidity and mortality in older adults: A systematic review and meta-analysis. Archives of Gerontology and Geriatrics. 2016;67:130–138. doi:10.1016/j.archger.2016.07.008

7. Makovski TT, Schmitz S, Zeegers MP, Stranges S, van den Akker M. Multimorbidity and quality of life: Systematic literature review and meta-analysis. Ageing Research Reviews. 2019;53:100903. doi:10.1016/j.arr.2019.04.005

8. Ryan A, Wallace E, O’Hara P, Smith SM. Multimorbidity and functional decline in community-dwelling adults: a systematic review. Health Qual Life Outcomes. 2015;13(1):168. doi:10.1186/s12955-015-0355-9

9. Dodds RM, Bunn JG, Hillman SJ, et al. Simple approaches to characterising multiple long-term conditions (multimorbidity) and rates of emergency hospital admission: Findings from 495,465 UK Biobank participants. Journal of Internal Medicine. 2023;293(1):100–109. doi:10.1111/joim.13567

10. Dodds RM, Granic A, Robinson SM, Sayer AA. Sarcopenia, long[term conditions, and multimorbidity: findings from UK Biobank participants. Journal of Cachexia, Sarcopenia and Muscle. 2020;11(1):62–68. doi:10.1002/jcsm.12503

11. Hurst C, Murray JC, Granic A, et al. Long-term conditions, multimorbidity, lifestyle factors and change in grip strength over 9 years of follow-up: Findings from 44,315 UK biobank participants. Age and Ageing. 2021;50(6):2222–2229. doi:10.1093/ageing/afab195

12. Hillman SJ, Dodds RM, Granic A, Witham MD, Sayer AA, Cooper R. Identifying combinations of long-term conditions associated with sarcopenia: a cross-sectional decision tree analysis in the UK Biobank study. BMJ Open. 2024;14(9):e085204. doi:10.1136/bmjopen-2024-085204

13. Cruz-Jentoft AJ, Bahat G, Bauer J, et al. Sarcopenia: revised European consensus on definition and diagnosis. Age Ageing. 2019;48(1):16–31. doi:10.1093/ageing/afy169

14. Cruz-Jentoft AJ, Sayer AA. Sarcopenia. The Lancet. 2019;393(10191):2636-2646. doi:10.1016/S0140-6736(19)31138-9

15. Sayer AA, Cooper R, Arai H, et al. Sarcopenia. Nat Rev Dis Primers. 2024;10(1):68. doi:10.1038/s41572-024-00550-w

16. Beaudart C, Demonceau C, Reginster JY, et al. Sarcopenia and health-related quality of life: A systematic review and meta-analysis. Journal of Cachexia, Sarcopenia and Muscle. 2023;14(3):1228–1243. doi:10.1002/jcsm.13243

17. Bricca A, Harris LK, Jager M, Smith SM, Juhl CB, Skou ST. Benefits and harms of exercise therapy in people with multimorbidity: A systematic review and meta-analysis of randomised controlled trials. Ageing Res Rev. 2020;63(101128963):101166. doi:10.1016/j.arr.2020.101166

18. Young HML, Henson J, Dempsey PC, et al. Physical activity and sedentary behaviour interventions for people living with both frailty and multiple long-term conditions and their informal carers: a scoping review and stakeholder consultation. Age and Ageing. 2024;53(11):afae255. doi:10.1093/ageing/afae255

19. Hurst C, Sayer AA. Improving muscle strength and physical function in older people living with sarcopenia and physical frailty: Not all exercise is created equal. Journal of the Royal College of Physicians of Edinburgh. Published online June 17, 2022:147827152211048. doi:10.1177/14782715221104859

20. Borde R, Hortobágyi T, Granacher U. Dose–Response Relationships of Resistance Training in Healthy Old Adults: A Systematic Review and Meta-Analysis. Sports Med. 2015;45(12):1693–1720. doi:10.1007/s40279-015-0385-9

21. Steib S, Schoene D, Pfeifer K. Dose–response relationship of resistance training in older adults: a meta-analysis. Br J Sports Med. 2010;42(5):902–914. doi:10.1136/bjsm.2010.083246

22. Dent E, Morley JE, Cruz-Jentoft AJ, et al. International Clinical Practice Guidelines for Sarcopenia (ICFSR): Screening, Diagnosis and Management. J Nutr Health Aging. 2018;22(10):1148–1161. doi:10.1007/s12603-018-1139-9

23. Hurst C, Robinson SM, Witham MD, et al. Resistance exercise as a treatment for sarcopenia: prescription and delivery. Age and Ageing. 2022;51(2):afac003. doi:10.1093/ageing/afac003

24. Cornelissen V, Hanssen H, Hurst C, et al. Influence of exercise and nutrition on sarcopenia in cardiovascular disease: a Scientific Statement of the European Association of Preventive Cardiology of the European Society of Cardiology. Eur J Prev Cardiol. Published online July 15, 2025:zwaf432. doi:10.1093/eurjpc/zwaf432

25. Pedersen BK, Saltin B. Exercise as medicine – evidence for prescribing exercise as therapy in 26 different chronic diseases. Scandinavian Journal of Medicine & Science in Sports. 2015;25(S3):1–72. doi:10.1111/sms.12581

26. Hurst C, Dismore L, Granic A, et al. Attitudes and barriers to resistance exercise training for older adults living with multiple long-term conditions, frailty, and a recent deterioration in health: qualitative findings from the Lifestyle in Later Life – Older People’s Medicine (LiLL-OPM) study. BMC Geriatr. 2023;23(1):772. doi:10.1186/s12877-023-04461-5

27. de Souto Barreto P. Exercise for Multimorbid Patients in Primary Care: One Prescription for All? Sports Med. 2017;47(11):2143–2153. doi:10.1007/s40279-017-0725-z

28. Hurst C, Dismore L, Granic A, et al. The feasibility and acceptability of engaging older adults living with multiple long-term conditions, frailty, and a recent deterioration in health in research: Findings from the Lifestyle in Later Life – Older People’s Medicine (LiLL-OPM) study. BMC Geriatr. 2024;24(1):831. doi:10.1186/s12877-024-05406-2

29. Yarnall AJ, Sayer AA, Clegg A, Rockwood K, Parker S, Hindle JV. New horizons in multimorbidity in older adults. Age and Ageing. 2017;46(6):882–888. doi:10.1093/ageing/afx150

30. Arksey H, O’Malley L. Scoping studies: towards a methodological framework. International Journal of Social Research Methodology. 2005;8(1):19–32. doi:10.1080/1364557032000119616

31. Tricco AC, Lillie E, Zarin W, et al. PRISMA Extension for Scoping Reviews (PRISMA-ScR): Checklist and Explanation. Ann Intern Med. 2018;169(7):467–473. doi:10.7326/M18-0850

32. Ho ISS, Azcoaga-Lorenzo A, Akbari A, et al. Examining variation in the measurement of multimorbidity in research: a systematic review of 566 studies. Lancet Public Health. 2021;6(8):e587–e597. doi:10.1016/S2468-2667(21)00107-9

33. Al-Mhanna SB, Batrakoulis A, Mohamed M, et al. Home-based circuit training improves blood lipid profile, liver function, musculoskeletal fitness, and health-related quality of life in overweight/obese older adult patients with knee osteoarthritis and type 2 diabetes: a randomized controlled trial during the COVID-19 pandemic. BMC Sports Sci Med Rehabil. 2024;16(1):125. doi:10.1186/s13102-024-00915-4

34. Barker K, Holland AE, Lee AL, et al. A rehabilitation programme for people with multimorbidity versus usual care: A pilot randomized controlled trial. Journal of Comorbidity. 2018;8(1):1–11. doi:10.1177/2235042X18783918

35. Baptista LC, Machado-Rodrigues AM, Veríssimo MT, Martins RA. Exercise training improves functional status in hypertensive older adults under angiotensin converting enzymes inhibitors medication. Experimental Gerontology. 2018;109:82–89. doi:10.1016/j.exger.2017.06.013

36. Bennett PN, Hussein WF, Matthews K, et al. An Exercise Program for Peritoneal Dialysis Patients in the United States: A Feasibility Study. Kidney Medicine. 2020;2(3):267–275. doi:10.1016/j.xkme.2020.01.005

37. Bernocchi P, Vitacca M, La Rovere MT, et al. Home-based telerehabilitation in older patients with chronic obstructive pulmonary disease and heart failure: a randomised controlled trial. Age and Ageing. 2018;47(1):82–88. doi:10.1093/ageing/afx146

38. Berry MJ, Rejeski WJ, Adair NE, Ettinger, WH, Zaccaro DJ, Sevick MA. A Randomized, Controlled Trial Comparing Long-term and Short-term Exercise in Patients With Chronic Obstructive Pulmonary Disease: Journal of Cardiopulmonary Rehabilitation. 2003;23(1):60–68. doi:10.1097/00008483-200301000-00011

39. Blasco-Lafarga C, Sanchis-Soler G, Llorens P. Multicomponent Physical Exercise Training in Multimorbid and Palliative Oldest Adults. IJERPH. 2021;18(17):8896. doi:10.3390/ijerph18178896

40. Bourne S, DeVos R, North M, et al. Online versus face-to-face pulmonary rehabilitation for patients with chronic obstructive pulmonary disease: randomised controlled trial. BMJ open. 2017;7(7):e014580. doi:10.1136/bmjopen-2016-014580

41. Buford TW, Manini TM, Hsu FC, et al. Angiotensin-converting enzyme inhibitor use by older adults is associated with greater functional responses to exercise. J Am Geriatr Soc. 2012;60(7):1244–1252. doi:10.1111/j.1532-5415.2012.04045.x

42. Celli A., Barnouin Y., Jiang B., et al. Lifestyle Intervention Strategy to Treat Diabetes in Older Adults: A Randomized Controlled Trial. Diabetes Care. 2022;45(9):1943–1952. doi:10.2337/dc22-0338

43. Chang CF, Lin KC, Chen WM, Jane SW, Yeh SH, Wang TJ. Effects of a Home-Based Resistance Training Program on Recovery From Total Hip Replacement Surgery: Feasibility and Pilot Testing. Journal of Nursing Research. 2017;25(1):21–30. doi:10.1097/jnr.0000000000000128

44. Charikiopoulou M, Nikolaidis PT, Knechtle B, Rosemann T, Rapti A, Trakada G. Subjective and Objective Outcomes in Patients With COPD After Pulmonary Rehabilitation – The Impact of Comorbidities. Front Physiol. 2019;10:286. doi:10.3389/fphys.2019.00286

45. Chen SM, Shen FC, Chen JF, Chang WD, Chang NJ. Effects of resistance exercise on glycated hemoglobin and functional performance in older patients with comorbid diabetes mellitus and knee osteoarthritis: A randomized trial. International Journal of Environmental Research and Public Health. 2020;17(1). doi:10.3390/ijerph17010224

46. Choi M, Kim JS, Park CY, Choi Y, Yoon T, Bae J. Feasibility of Whole-Body Resistance Training With Social Support Reinforcement for Older Adults Living Alone: A Mixed-Methods Pilot Study. Journal of Gerontological Nursing. 2024;50(10):34–41. doi:10.3928/00989134-20240913-01

47. Coca-Martinez M, Girsowicz E, Doonan RJ, et al. Multimodal Prehabilitation for Peripheral Arterial Disease Patients with Intermittent Claudication—A Pilot Randomized Controlled Trial. Annals of Vascular Surgery. 2024;107:2–12. doi:10.1016/j.avsg.2023.09.101

48. Dao E., Davis J.C., Sharma D., Chan A., Nagamatsu L.S., Liu-Ambrose T. Change in Body Fat Mass Is Independently Associated with Executive Functions in Older Women: A Secondary Analysis of a 12-Month Randomized Controlled Trial. PLoS ONE. 2013;8(1):e52831. doi:10.1371/journal.pone.0052831

49. Edelmann F, Gelbrich G, Düngen HD, et al. Exercise Training Improves Exercise Capacity and Diastolic Function in Patients With Heart Failure With Preserved Ejection Fraction. Journal of the American College of Cardiology. 2011;58(17):1780–1791. doi:10.1016/j.jacc.2011.06.054

50. Felcar JM, Probst VS, de Carvalho DR, et al. Effects of exercise training in water and on land in patients with COPD: a randomised clinical trial. Physiotherapy. 2018;104(4):408–416. doi:10.1016/j.physio.2017.10.009

51. Galvao DA, Spry N, Denham J, et al. A multicentre year-long randomised controlled trial of exercise training targeting physical functioning in men with prostate cancer previously treated with androgen suppression and radiation from TROG 03.04 RADAR. Eur Urol. 2014;65(5):856–864. doi:10.1016/j.eururo.2013.09.041

52. Gary RA, Cress ME, Higgins MK, Smith AL, Dunbar SB. Combined Aerobic and Resistance Exercise Program Improves Task Performance in Patients With Heart Failure. Archives of Physical Medicine & Rehabilitation. 2011;92(9):1371–1381. doi:10.1016/j.apmr.2011.02.022

53. Genest F, Lindström S, Scherer S, Schneider M, Seefried L. Feasibility of simple exercise interventions for men with osteoporosis – A prospective randomized controlled pilot study. Bone Reports. 2021;15:101099. doi:10.1016/j.bonr.2021.101099

54. GilHerrero L, Courneya KS, McNeely ML, et al. Effects of a Clinical Exercise Program on Health-Related Fitness and Quality of Life in Spanish Cancer Patients Receiving Adjuvant Therapy. Integrative Cancer Therapies. 2022;21. doi:10.1177/15347354221141715

55. Hall KS, Morey MC, Beckham JC, et al. Warrior Wellness: A Randomized Controlled Pilot Trial of the Effects of Exercise on Physical Function and Clinical Health Risk Factors in Older Military Veterans With PTSD. J Gerontol A Biol Sci Med Sci. 2020;75(11):2130–2138. doi:10.1093/gerona/glz255

56. Hellberg M, Höglund P, Svensson P, Clyne N. Randomized Controlled Trial of Exercise in CKD—The RENEXC Study. Kidney International Reports. 2019;4(7):963–976. doi:10.1016/j.ekir.2019.04.001

57. Hetherington S, Henwood T, Swinton P, et al. Engineering Improved Balance Confidence in Older Adults With Complex Health Care Needs: Learning From the Muscling Up Against Disability Study. Archives of Physical Medicine & Rehabilitation. 2018;99(8):1525–1532. doi:10.1016/j.apmr.2018.03.004

58. Hinrichs T, Bücker B, Klaaßen[Mielke R, et al. Home[Based Exercise Supported by General Practitioner Practices: Ineffective in a Sample of Chronically Ill, Mobility[Limited Older Adults (the HOME fit Randomized Controlled Trial). J American Geriatrics Society. 2016;64(11):2270–2279. doi:10.1111/jgs.14392

59. Hjermundrud V, Hilding GF, Gjøvaag T. Four weeks of inpatient comprehensive prosthetic rehabilitation achieves contrasting results in different groups of prosthetic users. Prosthetics and Orthotics International. 2024;48(6):634. doi:10.1097/PXR.0000000000000324

60. Houben LHP, Overkamp M, Senden JMG, et al. Benefits of resistance training are not preserved after cessation of supervised training in prostate cancer patients on androgen deprivation therapy. European Journal of Sport Science. 2024;24(1):116–126. doi:10.1002/ejsc.12050

61. Jacobs JL, Marcus RL, Morrell G, LaStayo P. Resistance exercise with older fallers: its impact on intermuscular adipose tissue. Biomed Res Int. 2014;2014(101600173):398960. doi:10.1155/2014/398960

62. Kaye DR, Schafer C, Thelen-Perry S, et al. The Feasibility and Impact of a Presurgical Exercise Intervention Program (Prehabilitation) for Patients Undergoing Cystectomy for Bladder Cancer. Urology. 2020;145:106–112. doi:10.1016/j.urology.2020.05.104

63. Kelly MA, Finley M, Lichtman SW, Hyland MR, Edeer AO. Comparative Analysis of High-Velocity Versus Low-Velocity Exercise on Outcomes After Total Knee Arthroplasty: A Randomized Clinical Trial. Journal of Geriatric Physical Therapy. 2016;39(4):178–189. doi:10.1519/JPT.0000000000000070

64. Kitzman DW, Whellan DJ, Duncan P, et al. Physical Rehabilitation for Older Patients Hospitalized for Heart Failure. N Engl J Med. 2021;385(3):203–216. doi:10.1056/NEJMoa2026141

65. Krumov J, Obretenov V, Bozov H, et al. Is group-based physical therapy superior to individual rehabilitation in elderly adults after total knee arthroplasty? A prospective observational study. European Journal of Translational Myology. 2022;32(4). doi:10.4081/ejtm.2022.10984

66. Lauze M, Martel DD, Aubertin-Leheudre M. Feasibility and Effects of a Physical Activity Program Using Gerontechnology in Assisted Living Communities for Older Adults. J AM MED DIR ASSOC. 2017;18(12):1069–1075. doi:10.1016/j.jamda.2017.06.030

67. Layne JE, Arabelovic S, Wilson LB, et al. Community-Based Strength Training Improves Physical Function in Older Women With Arthritis. American Journal of Lifestyle Medicine. 2009;3(6):466–473. doi:10.1177/1559827609342061

68. Lee WJ, Peng LN, Lin MH, Lin CH, Chen LK. Clinical Efficacy of Multidomain Interventions among Multimorbid Older People Stratified by the Status of Physio-Cognitive Declines: A Secondary Analysis from the Randomized Controlled Trial for Healthy Aging. Journal of Nutrition, Health & Aging. 2022;26(10):909–917. doi:10.1007/s12603-022-1843-3

69. Legault EP, Ribeiro PAB, Moreau-Amaru D, et al. The PREPARE Study: Acceptability and Feasibility of a Telehealth Trimodal Prehabilitation Program for Women with Endometrial Neoplasia. Current Oncology. 2025;32(1):55. doi:10.3390/curroncol32010055

70. Mangani I, Cesari M, Kritchevsky SB, et al. Physical exercise and comorbidity. Results from the Fitness and Arthritis in Seniors Trial (FAST). Aging Clinical and Experimental Research. 2006;18(5):374–380. doi:10.1007/BF03324833

71. Martin-Alemany G., Perez-Navarro M., Wilund K.R., et al. Effect of Intradialytic Oral Nutritional Supplementation with or without Exercise Improves Muscle Mass Quality and Physical Function in Hemodialysis Patients: A Pilot Study. Nutrients. 2022;14(14):2946. doi:10.3390/nu14142946

72. Merchant RA, Chan YH, Hui RJY, et al. Possible Sarcopenia and Impact of Dual-Task Exercise on Gait Speed, Handgrip Strength, Falls, and Perceived Health. Frontiers in Medicine. 2021;8. doi:10.3389/fmed.2021.660463

73. Mulrow CD, Gerety MB, Kanten D, et al. A Randomized Trial of Physical Rehabilitation for Very Frail Nursing Home Residents. JAMA: The Journal of the American Medical Association. 1994;271(7):519–524. doi:10.1001/jama.1994.03510310049037

74. Nemoto M, Nemoto K, Sasai H, Higashi S, Ota M, Arai T. Long-Term Multimodal Exercise Intervention for Patients with Frontotemporal Lobar Degeneration: Feasibility and Preliminary Outcomes. Dement Geriatr Cogn Disord Extra. 2024;15(1):19–29. doi:10.1159/000542994

75. O’Brien KK, Davis AM, Chan Carusone S, et al. Examining the impact of a community-based exercise intervention on cardiorespiratory fitness, cardiovascular health, strength, flexibility and physical activity among adults living with HIV: A three-phased intervention study. PLoS ONE. 2021;16(9):e0257639. doi:10.1371/journal.pone.0257639

76. Oh, Kim D.-Y., Bae J.-H., Lim J.-Y. Effects of rural community-based integrated exercise and health education programs on the mobility function of older adults with knee osteoarthritis. Aging Clin Exp Res. 2021;33(11):3005–3014. doi:10.1007/s40520-020-01474-7

77. Ory MG, Smith ML, Luohua Jiang, et al. Texercise Effectiveness: Impacts on Physical Functioning and Quality of Life. Journal of Aging & Physical Activity. 2015;23(4):622–629. doi:10.1123/japa.2014-0072

78. Pizarro-Mena R, Duran-Aguero S, Parra-Soto S, Vargas-Silva F, Bello-Lepe S, Fuentes-Alburquenque M. Effects of a Structured Multicomponent Physical Exercise Intervention on Quality of Life and Biopsychosocial Health among Chilean Older Adults from the Community with Controlled Multimorbidity: A Pre-Post Design. Int J Environ Res Public Health. 2022;19(23):15842. doi:10.3390/ijerph192315842

79. Potiaumpai M, Caru M, Mineishi S, Naik S, Zemel BS, Schmitz KH. IMPROVE-BMT: A Pilot Randomized Controlled Trial of Prehabilitation Exercise for Adult Hematopoietic Stem Cell Transplant Recipients. Journal of Clinical Medicine. 2024;13(7):2052. doi:10.3390/jcm13072052

80. Rauzi MR, Abbate LM, Churchill L, et al. Multicomponent telerehabilitation program for veterans with multimorbidity: A randomized controlled feasibility study. PM&R. 2025;17(5):548–562. doi:10.1002/pmrj.13299

81. Reeves GR, Whellan DJ, O’Connor CM, et al. A Novel Rehabilitation Intervention for Older Patients With Acute Decompensated Heart Failure: The REHAB-HF Pilot Study. JACC Heart Fail. 2017;5(5):359–366. doi:10.1016/j.jchf.2016.12.019

82. Rimmer JH, Riley B, Creviston T, Nicola T. Exercise training in a predominantly African-American group of stroke survivors. Medicine and Science in Sports and Exercise. 2000;32(12):1990–1996. doi:10.1097/00005768-200012000-00004

83. Rimmer JH, Nicola T, Riley B, Creviston T. Exercise training for African Americans with disabilities residing in difficult social environments. American Journal of Preventive Medicine. 2002;23(4):290–295. doi:10.1016/S0749-3797(02)00517-2

84. Rodriguez Mañas L, Laosa O, Vellas B, et al. Effectiveness of a multimodal intervention in functionally impaired older people with type 2 diabetes mellitus. J cachexia sarcopenia muscle. 2019;10(4):721–733. doi:10.1002/jcsm.12432

85. Ryan A, Smith SM, Cummins V, Murphy C, Galvin R. Development and feasibility of an inter-agency physical activity and education programme for adults with multimorbidity in primary care: Activ8. Journal of Multimorbidity & Comorbidity. 2022;12:1–13. doi:10.1177/26335565221142350

86. Seino S, Nishi M, Murayama H, et al. Effects of a multifactorial intervention comprising resistance exercise, nutritional and psychosocial programs on frailty and functional health in community-dwelling older adults: A randomized, controlled, cross-over trial. Geriatrics and Gerontology International. 2017;17(11):2034–2045. doi:10.1111/ggi.13016

87. Serra-Prat M, Sist X, Domenich R, et al. Effectiveness of an intervention to prevent frailty in pre-frail community-dwelling older people consulting in primary care: A randomised controlled trial. Age and Ageing. 2017;46(3):401–407. doi:10.1093/ageing/afw242

88. Servantes DM, Pelcerman A, Salvetti XM, et al. Effects of home-based exercise training for patients with chronic heart failure and sleep apnoea: a randomized comparison of two different programmes. Clin Rehabil. 2012;26(1):45–57. doi:10.1177/0269215511403941

89. Shiozaki K, Asaeda M, Hashimoto T, et al. Effects of Physiatrist and Physiotherapist-supervised Therapeutic Exercise on Physical Function in Frail Older Patients with Multimorbidity. Progress in rehabilitation medicine. 2023;8:20230012–20230012. doi:10.2490/prm.20230012

90. Soria-Comes T, Climent-Gregori M, Maestu-Maiques I, et al. Effect of a Physical Exercise Intervention on Physical Function Parameters and Blood Analytical Changes in Lung Cancer Survivors: A Feasibility Study. Clinics and Practice. 2024;14(5):2202–2216. doi:10.3390/clinpract14050173

91. Stevens-Lapsley JE, Derlein D, Churchill L, et al. High-intensity home health physical therapy among older adult Veterans: A randomized controlled trial. Journal of the American Geriatrics Society. Published online 2023. doi:10.1111/jgs.18413

92. Tao X, Chow SK, Wong FK. A nurse-led case management program on home exercise training for hemodialysis patients: A randomized controlled trial. International Journal of Nursing Studies. 2015;52(6):1029–1041. doi:10.1016/j.ijnurstu.2015.03.013

93. Teuwen MMH, van Weely SFE, Vliet Vlieland TPM, et al. Effectiveness of longstanding exercise therapy compared with usual care for people with rheumatoid arthritis and severe functional limitations: a randomised controlled trial. Annals of the Rheumatic Diseases. 2024;83(4):437–445. doi:10.1136/ard-2023-224912

94. Tikkanen P, Lonnroos E, Sipila S, Nykanen I, Sulkava R, Hartikainen S. Effects of comprehensive health assessment and targeted intervention on chair rise capacity in active and inactive community-dwelling older people. Gerontology. 2013;59(4):324–327. doi:10.1159/000347197

95. Ullrich P, Werner C, Schonstein A, et al. Effects of a Home-Based Physical Training and Activity Promotion Program in Community-Dwelling Older Persons with Cognitive Impairment after Discharge from Rehabilitation: A Randomized Controlled Trial. J Gerontol A Biol Sci Med Sci. 2022;77(12):2435–2444. doi:10.1093/gerona/glac005

96. van den Helder J, Mehra S, van Dronkelaar C, et al. Blended home-based exercise and dietary protein in community-dwelling older adults: a cluster randomized controlled trial. Journal of Cachexia, Sarcopenia and Muscle. 2020;11(6):1590–1602. doi:10.1002/jcsm.12634

97. Weiner D.E., Liu C.K., Miao S., et al. Effect of Long-term Exercise Training on Physical Performance and Cardiorespiratory Function in Adults With CKD: A Randomized Controlled Trial. Am J Kidney Dis. 2023;81(1):59–66. doi:10.1053/j.ajkd.2022.06.008

98. Wilson RL, Newton RU, Taaffe DR, Hart NH, Lyons-Wall P, Galvao DA. Weight Loss for Obese Prostate Cancer Patients on Androgen Deprivation Therapy. Med Sci Sports Exerc. 2021;53(3):470–478. doi:10.1249/MSS.0000000000002509

99. Winters-Stone KM, Horak F, Dieckmann NF, et al. GET FIT: A Randomized Clinical Trial of Tai Ji Quan Versus Strength Training for Fall Prevention After Chemotherapy in Older, Postmenopausal Women Cancer Survivors. Journal of Clinical Oncology. 2023;41(18):3384–3396. doi:10.1200/JCO.22.01519

100. Zhou Y, Hellberg M, Hellmark T, Höglund P, Clyne N. Muscle mass and plasma myostatin after exercise training: A substudy of Renal Exercise (RENEXC) — A randomized controlled trial. Nephrology Dialysis Transplantation. 2021;36(1):95–103. doi:10.1093/NDT/GFZ210

101. de Vries NM, van Ravensberg CD, Hobbelen JSM, Olde Rikkert MGM, Staal JB, Nijhuis-van der Sanden MWG. Effects of physical exercise therapy on mobility, physical functioning, physical activity and quality of life in community-dwelling older adults with impaired mobility, physical disability and/or multi-morbidity: A meta-analysis. Ageing Research Reviews. 2012;11(1):136–149. doi:10.1016/j.arr.2011.11.002

102. Forsyth F, Soh CL, Elks N, et al. Exercise Modalities in Multi-Component Interventions for Older adults with Multi-Morbidity: A Systematic Review and Narrative Synthesis. J Frailty Aging. Published online March 26, 2024. doi:10.14283/jfa.2024.28

103. Fuller JT, Hartland MC, Maloney LT, Davison K. Therapeutic effects of aerobic and resistance exercises for cancer survivors: a systematic review of meta-analyses of clinical trials. Br J Sports Med. 2018;52(20):1311–1311. doi:10.1136/bjsports-2017-098285

104. Fisher S, Smart NA, Pearson MJ. Resistance training in heart failure patients: a systematic review and meta-analysis. Heart Fail Rev. 2022;27(5):1665–1682. doi:10.1007/s10741-021-10169-8

105. Fisher JP, Steele J, Gentil P, Giessing J, Westcott WL. A minimal dose approach to resistance training for the older adult; the prophylactic for aging. Experimental Gerontology. 2017;99:80–86. doi:10.1016/j.exger.2017.09.012

106. Brightwell CR, Markofski MM, Moro T, et al. Moderate-intensity aerobic exercise improves skeletal muscle quality in older adults. TRANSLATIONAL SPORTS MEDICINE. 2019;2(3):109–119. doi:10.1002/tsm2.70

107. Harber MP, Konopka AR, Douglass MD, et al. Aerobic exercise training improves whole muscle and single myofiber size and function in older women. American Journal of Physiology-Regulatory, Integrative and Comparative Physiology. 2009;297(5):R1452–R1459. doi:10.1152/ajpregu.00354.2009

108. Caulfield L, Arnold S, Biase SD, et al. The Benchmarking Exercise Programme for Older People (BEPOP): Design, Results and Recommendations from The First Wave of Data Collection. JFSF. 2024;9(3):169–183. doi:10.22540/JFSF-09-169

109. Dekker J, Buurman BM, van der Leeden M. Exercise in people with comorbidity or multimorbidity. Health Psychology. 2019;38(9):822–830. doi:10.1037/hea0000750

110. Slade SC, Dionne CE, Underwood M, Buchbinder R. Consensus on Exercise Reporting Template (CERT): Explanation and Elaboration Statement. Br J Sports Med. 2016;50(23):1428–1437. doi:10.1136/bjsports-2016-096651

111. Taylor KL, Weston M, Batterham AM. Evaluating Intervention Fidelity: An Example from a High-Intensity Interval Training Study. Piacentini MF, ed. PLoS ONE. 2015;10(4):e0125166. doi:10.1371/journal.pone.0125166

112. Soysal P, Hurst C, Demurtas J, et al. Handgrip strength and health outcomes: Umbrella review of systematic reviews with meta-analyses of observational studies. Journal of Sport and Health Science. 2021;10(3):290–295. doi:10.1016/j.jshs.2020.06.009

113. Tieland M, Verdijk LB, de Groot LCPGM, van Loon LJC. Handgrip strength does not represent an appropriate measure to evaluate changes in muscle strength during an exercise intervention program in frail older people. Int J Sport Nutr Exerc Metab. 2015;25(1):27–36. doi:10.1123/ijsnem.2013-0123

114. van der Leeden M, Stuiver MM, Huijsmans R, Geleijn E, de Rooij M, Dekker J. Structured clinical reasoning for exercise prescription in patients with comorbidity. Disability and Rehabilitation. 2020;42(10):1474–1479. doi:10.1080/09638288.2018.1527953

115. Cooper R, Bunn JG, Richardson SJ, et al. Rising to the challenge of defining and operationalising multimorbidity in a UK hospital setting: the ADMISSION research collaborative. Eur Geriatr Med. Published online March 6, 2024. doi:10.1007/s41999-024-00953-8

