## Supporting information for "Resistance exercise training for skeletal muscle health in adults living with multiple long-term conditions: a scoping review"

**Contents**


Table S1: Full search strategy

Table S2: Characteristics of included studies

Table S3: Exercise intervention details

Table S4: Study outcomes

Table S5: Preferred Reporting Items for Systematic reviews and Meta-Analyses extension for Scoping Reviews (PRISMA-ScR) Checklist

**Table S1: Full search strategy**

| *Database searching* | |
|---|---|
| Database | Search strategy |
| Ovid interface<br>**MEDLINE**<br>**Embase** | 1. exp Comorbidity/<br>2. (comorbidit* or multimorbid* or polymorbidit* or multiple LTC* or multi?condition* or multiple chronic condition* or multimorbidity).ti,ab,kf.<br>3. ((multiple or coexisting or co-existing or concurrent or con-current or comorbid or co-morbid) adj2 (disease* or illness* or condition* or diagnos* or morbid*)).ti,ab,kf.<br>4. 1 OR 2 OR 3<br>5. exp Exercise/<br>6. exp Exercise Therapy/<br>7. (exercise or strength training or resistance training or weight training or weight lifting or resistance exercise or muscle-strengthening exercise or theraband or aerobic training or endurance training or combined training or concurrent training or combined exercise or concurrent exercise).ti,ab,kf.<br>8. 5 OR 6 OR 7<br>9. exp Muscle, Skeletal/ or muscle size.mp.<br>10. muscle mass.mp.<br>11. (muscle mass or skeletal muscle mass or skeletal muscle index or total muscle mass or appendicular muscle mass or appendicular lean mass or muscle size or lean mass).ti,ab,kf<br>12. 9 or 10 or 11<br>13. muscle strength.mp. or exp Muscle Strength/<br>14. (muscle strength or muscle function or grip strength or muscle power or leg strength or knee extensor strength).ti,ab,kf.<br>15. 13 OR 14<br>16. exp Physical Functional Performance/<br>17. ((walking speed or gait or gait speed or chair rises or chair stands or physical performance or physical function or "Timed Up and Go")).ti,ab,kf.<br>18. 15 or 16<br>19. 12 OR 15 OR 18<br>20. 4 AND 8 AND 19<br>21. Limit 20 to (english language and humans) |
| EBSCO interface<br>**CINAHL** | 1. comorbidit* or multimorbidity* or polymorbidit* or multiple LTC* or multi?condition* or multiple chronic condition<br>2. ((multiple or coexisting or co-existing or concurrent or con-current or comorbid or co-morbid) n2 (disease* or illness* or condition* or diagnos* or morbid*))<br>3. 1 OR 2<br>4. exercise or strength training or resistance training or weight training or weight lifting or resistance exercise or muscle-strengthening exercise or theraband or aerobic training or endurance training or combined training or concurrent training or combined exercise or concurrent exercise<br>5. muscle mass or skeletal muscle mass or skeletal muscle index or total muscle mass or appendicular muscle mass or appendicular lean mass or muscle size or lean mass<br>6. muscle strength or muscle function or grip strength or muscle power or leg strength or knee extensor strength |

| | |
|---|---|
| | 7. walking speed or gait or gait speed or chair rises or chair stands or physical performance or physical function or Timed Up and Go<br>8. 5 OR 6 OR 7<br>9. 3 AND 4 AND 8<br>Limiters – English Language; Peer reviewed |
| **Scopus** | 1. TITLE-ABS-KEY ( ( comorbidit* OR multimorbid* OR polymorbidit* OR "multiple ltc*" OR multi?condition* OR "multiple chronic condition*" OR multimorbidity ) )<br>2. TITLE-ABS-KEY ( ( multiple OR coexisting OR co-existing OR concurrent OR con-current OR comorbid OR co-morbid ) W/2 ( disease* OR illness* OR condition* OR diagnos* OR morbid* ) )<br>3. 1 OR 2<br>4. TITLE-ABS-KEY ( exercise OR "strength training" OR "resistance training" OR "weight training" OR "weight lifting" OR "resistance exercise" OR "muscle-strengthening exercise" OR "theraband" OR "aerobic training" OR "endurance training" OR "combined training" OR "concurrent training" OR "combined exercise" OR "concurrent exercise" )<br>5. TITLE-ABS-KEY ( "muscle mass" OR "skeletal muscle mass" OR "skeletal muscle index" OR "total muscle mass" OR "appendicular muscle mass" OR "appendicular lean mass" OR "muscle size" OR "lean mass" )<br>6. TITLE-ABS-KEY ( "muscle strength" OR "muscle function" OR "grip strength" OR "muscle power" OR "leg strength" OR "knee extensor strength" )<br>7. TITLE-ABS-KEY ( "walking speed" OR "gait" OR "gait speed" OR "chair rises" OR "chair stands" OR "physical performance" OR "physical function" OR "Timed Up and Go" )<br>8. #5 OR #6 OR #7<br>9. #3 AND #4 AND #8<br>10. Limit to: English Language |
| **Web of Science** (ALL databases: Web of Science Core Collection, KCI-Korean Journal Database, Russian Science Citation Index, SciELO Citation Index, Zoological Record) | 1. comorbidity (Topic) or multimorbidity (Topic) or comorbidit* OR multimorbid* OR polymorbidit* OR "multiple ltc*" OR multi?condition* OR "multiple chronic condition**"** (Topic)<br>2. ( exercise OR "strength training" OR "resistance training" OR "weight training" OR "weight lifting" OR "resistance exercise" OR "muscle-strengthening exercise" OR "theraband" OR "aerobic training" OR "endurance training" OR "combined training" OR "concurrent training" OR "combined exercise" OR "concurrent exercise" ) (Topic)<br>3. ( "muscle mass" OR "skeletal muscle mass" OR "skeletal muscle index" OR "total muscle mass" OR "appendicular muscle mass" OR "appendicular lean mass" OR "muscle size" OR "lean mass" ) (Topic) or ( "muscle strength" OR "muscle function" OR "grip strength" OR "muscle power" OR "leg strength" OR "knee extensor strength" ) (Topic) or ( "walking speed" OR "gait" OR "gait speed" OR "chair rises" OR "chair stands" OR "physical performance" OR "physical function" OR "Timed Up and Go" ) (Topic)<br>4. #1 AND #2 AND #3 and English (Languages) |
| **Cochrane Central Register of Controlled** | 1. MeSH descriptor: [Comorbidity] explode all trees<br>2. comorbidit* or multimorbidity* or polymorbidit* or "multiple LTC*" or multi?condition* or "multiple chronic condition*" or multimorbidity |

| | |
|---|---|
| **Trials (CENTRAL)** | 3. ((multiple or coexisting or co-existing or concurrent or con-current or comorbid or co-morbid) NEAR/2 (disease* or illness* or condition* or diagnos* or morbid*))<br>4. #1 OR #2 OR #3<br>5. MeSH descriptor: [Exercise] explode all trees<br>6. MeSH descriptor: [Exercise Therapy] explode all trees<br>7. (exercise or "strength training" or "resistance training" or "weight training" or "weight lifting" or "resistance exercise" or "muscle-strengthening exercise" or theraband or "aerobic training" or "endurance training" or "combined training" or "concurrent training" or "combined exercise" or "concurrent exercise")<br>8. #5 OR #6 OR #7<br>9. MeSH descriptor: [Muscles] explode all trees<br>10. muscle mass or skeletal muscle mass or skeletal muscle index or total muscle mass or appendicular muscle mass or appendicular lean mass or muscle size or lean mass<br>11. MeSH descriptor: [Muscle Strength] explode all trees<br>12. "muscle strength" or "muscle function" or "grip strength" or "muscle power" or "leg strength" or "knee extensor strength"<br>13. MeSH descriptor: [Physical Functional Performance] explode all trees<br>14. "walking speed" or "gait" or "gait speed" or "chair rises" or "chair stands" or "physical performance" or "physical function" or "Timed Up and Go"<br>15. #9 OR #10 OR #11 OR #12 OR #13 OR #14<br>16. #4 AND #8 AND #15 in Trials |

*Registry searching*

| Registry | Search strategy |
|---|---|
| ISRCTN (https://www.isrctn.com) | Text search: exercise<br>AND<br>Condition: multimorbidity |
| ClinicalTrials.gov (https://clinicaltrials.gov) | Condition or disease: multimorbidity<br>AND<br>Other terms: exercise |

**Table S2: Characteristics of included studies (ordered alphabetically by first author surname)**

| | | | Participant characteristics | | | | Characterisation of MLTC | |
|---|---|---|---|---|---|---|---|---|
| Study, year | Country | Study design | Sample size (*n*) | Age (years)* | % female | Study population | Method used for identification of MLTC (definition) | Method used for characterising MLTC (e.g., how many conditions considered) |
| Al-Mhanna et al. 2024 [33] | Malaysia | RCT | 70 | 62 ± 6 | 56 | Overweight or obese older adult patients with knee OA and T2DM | T2DM and OA | N/A |
| Barker et al. 2018 [34] | Australia | Pilot RCT | 16 | 69 ± 9 | 37 | Patients with chronic disease | No. of conditions | Total of 40 conditions |
| Baptista et al. 2018 [35] | Portugal | Parallel, non-randomised | 418 | 67 ± 6 | 76 | Hypertensive older adults | No. of conditions | Not reported |
| Bennett et al. 2020 [36] | USA | Parallel randomized controlled feasibility study | 26 | 58 ± 17 | 46 | Peritoneal Dialysis Patients | Index condition (Kidney disease) + comorbid conditions | Not reported |
| Bernocchi et al. 2018 [37] | Italy | Randomised, open, controlled, multicentre trial | 112 | 70 ± 9 | 18 | Patients with combined COPD and CHF | No. of conditions (Coexistence of COPD and CHF) | Total of 2 conditions |
| Berry et al. 2003 [38] | USA | RCT | 140 | 68 (SD N/A) | 44 | Patients with COPD | No. of conditions (Index condition COPD + at least 1 additional condition) | Total of 7 conditions / List of 6 other conditions |
| Blasco-Lafarga et al. 2021 [39] | Spain | Single group, pre-post | 33 | 82 ± 6 | 45 | Multimorbid and Palliative adults | No. of conditions | Not reported |
| Bourne et al. 2017 [40] | UK | Parallel single-blind RCT | 90 | 70 (SD N/A) | 34 | Patients with Chronic Obstructive Pulmonary Disease | No. of conditions (Index condition COPD + at least 1 additional condition) | COPD + List of 12 categories of conditions |
| Buford et al. 2012 [41] | USA | RCT | 424 | 77 ± 4 | 69 | Older adults at risk for mobility disability | No. of conditions | Not reported |
| Celli et al. 2022 [42] | USA | RCT | 100 | HL: 71.4 ± 3.7, ILI: 72.3 ± 4.0 | 35 | Older adults with diabetes | No. of conditions (Index condition (Diabetes) + at least 1 additional condition) | Not reported |
| Chang et al. 2017 [43] | Taiwan | Single group pre-post | 30 | 68 ± 8 | 70 | Patients with osteoarthritis | No. of conditions (index condition osteoarthritis + at least 1 additional condition) | Not reported |

| | | | | | | | | |
|---|---|---|---|---|---|---|---|---|
| Charikiopoulou et al. 2019 [44] | Greece | Parallel groups, non-randomised | 32 | 66 ± 6 | 22 | Patients with COPD | No. of conditions (index condition COPD + at least 1 additional condition) | Not reported |
| Chen et al. 2020 [45] | Taiwan | Parallel groups, pre-post | 70 | Group 1: 66 ± 3; Group 2: 65 ± 3 | 51 | Patients with comorbid diabetes mellitus and knee osteoarthritis | T2DM and OA | N/A |
| Choi et al. 2024 [46] | Republic of Korea | Single group pre-post | 7 | 73 ± 5 | 71 | Older adults living alone | No. of conditions | Not reported |
| Coca-Martinez et al. 2024 [47] | Canada | Pilot RCT | 24 | Intervention: 72 ± 7; Control: 70 ± 8 | 58 | Peripheral arterial disease patients with intermittent claudication | No. of conditions (index condition PAD + at least 1 additional condition) | Not reported |
| Dao et al. 2013 [48] | Canada | Parallel groups, pre-post | 114 | 69 ± 3 | 100 | Community-dwelling older women | No. of conditions | Functional comorbidity index |
| Edelmann et al. 2011 [49] | Germany | RCT | 64 | 65 ± 7 | 56 | Patients with heart failure | No. of conditions (index condition heart failure + at least 1 additional condition) | Not reported |
| Felcar et al. 2018 [50] | Brazil | RCT | 36 | 69 | 36 | Patients with COPD | No. of conditions (index condition COPD + 1 additional condition) | Not reported |
| Galvao et al. 2014 [51] | Australia & New Zealand | RCT | 100 | 72 | 0 | Prostate cancer survivors | No. of conditions (index condition prostate cancer + 1 additional condition) | Not reported |
| Gary et al. 2011 [52] | USA | RCT | 24 | 60 ± 10 | 50 | Patients with heart failure | No. of conditions (index condition heart failure + 1 additional condition) | Not reported |
| Genest et al. 2021 [53] | Germany | Pilot RCT | 47 | 77 ± 6 | 0 | Men at risk of osteoporosis | No. of conditions (3 or more conditions) | Not reported |
| GilHerrero et al. 2022 [54] | Spain | Single group pre-post | 85 | 49 ± 11 | 92 | Cancer patients | Number of conditions (Cancer + additional conditions) | Not reported |
| Hall et al. 2020 [55] | USA | Pilot RCT | 54 | 67 | 9 | Older veterans with PTSD. | Number of conditions | Not reported |
| Hellberg et al. 2019 [56] | Sweden | RCT | 151 | 66 ± 14 | 65 | Patients with Chronic Kidney Disease | No. of conditions (index condition CKD + 1 additional condition) | Davies Comorbidity Score |
| Hetherington et al. 2018 [57] | Australia | RCT | 245 | 79 ± 6 | 78 | Older adults with complex care needs | No. of conditions | Not reported |

| | | | | | | | | |
|---|---|---|---|---|---|---|---|---|
| Hinrichs et al. 2016 [58] | Germany | RCT | 209 | 80 ± 5 | 74 | Chronically ill and mobility-limited older adults | No. of conditions (presence of at least 2 medical conditions) | Total of 11 conditions |
| Hjermundrud et al. 2024 [59] | Norway | Parallel groups | 64 | Exp users: 67 ± 11<br>New users: 65 ± 11<br>Con: 62 ± 7 | 36 | Adults with lower limb loss | No. of conditions | Not reported |
| Houben et al. 2024 [60] | Netherlands | RCT | 68 | 71 ± 6 | 0 | Prostate cancer patients | No. of conditions (index condition prostate cancer + at least 1 additional condition) | Not reported |
| Jacobs et al. 2014 [61] | USA | Randomised, parallel groups trial | 77 | 76 ± 7 | 73 | Older adults with multiple comorbidities and a history of falling | No. of conditions | Not reported |
| Kaye et al. 2020 [62] | USA | Single group pre-post | 54 | 71 ± 6 | 24 | Patients with biopsy-proven bladder cancer | No. of conditions | Not reported |
| Kelly et al. 2016 [63] | USA | Randomised, parallel groups trial | 38 | 71 ± 7 | 63 | Adults who had undergone primary unilateral total knee arthroplasty | No. of conditions | Not reported |
| Kitzman et al. 2021 [64] | USA | RCT | 349 | 73 ± 8 | 52 | Heart failure patients | No. of conditions | Not reported |
| Krumov et al. 2022 [65] | Bulgaria | Parallel groups trial | 130 | 72 ± 1 | 52 | Older adults following total knee arthroplasty | No. of conditions | Not reported |
| Lauze et al. 2017 [66] | Canada | RCT | 32 | Ex group: 80 ± 8<br>Con: 83 ± 7 | 76 | Older adults in assisted living communities | No. of conditions (presence of 2 or more medical conditions) | Not reported |
| Layne et al. 2009 [67] | USA | Pilot RCT | 33 | 73 ± 9 | 100 | Community-dwelling older women with OA | No. of conditions (index condition OA + at least 1 additional condition) | Not reported |
| Lee et al. 2022 [68] | Taiwan | RCT | 340 | 72 ± 6 | 58 | Community-living multimorbid older people | Defined by authors as multimorbidity (No. of conditions) | Not reported |
| Legault et al. 2025 [69] | Canada | Parallel groups | 39 | 63 ± 9 | 100 | Women with endometrial neoplasia | No. of conditions (index condition endometrial cancer + at least 1 additional condition) | Not reported |
| Mangani et al. 2006 [70] | USA | RCT | 435 | 69 ± 6 | 70 | Community dwelling individuals with knee OA | No. of conditions (index condition OA + at least 1 additional condition) | Total of 10 conditions |

| | | | | | | | | |
|---|---|---|---|---|---|---|---|---|
| Martin-Alemany et al. 2022 [71] | Mexico | Parallel groups trial | 24 | Nutrition: 38 ± 12 exercise: 29 ± 10 | 58 | Haemodialysis patients | No. of conditions (index condition CKD + at least 1 additional condition) | Not reported |
| Merchant et al. 2021 [72] | Singapore | Parallel groups trial | 151 | Exercise: 76 ± 7 Control: 73 ± 6 | 66 | Community-dwelling older adults | No. of conditions (presence of 2 or more medical conditions) | Total of 7 conditions |
| Mulrow et al. 1994 [73] | USA | RCT | 194 | 80 ± 8 | 71 | Frail nursing home residents | No. of conditions (presence of 2 or more medical conditions) | Total of 7 conditions |
| Nemoto et al. 2024 [74] | Japan | Single group, pre-post | 4 | 69 ± 10 | 0 | Patients with frontotemporal lobar degeneration | No. of conditions | Not reported |
| O'Brien et al. 2021 [75] | Canada | Single group pre-post | 108 | Median age 51 (IQR 45-59) | 11 | Community dwelling adults living with HIV | No. of conditions (index condition HIV + at least 1 additional condition) | Total of 5 conditions (only the most common, >30%, were reported) |
| Oh et al. 2021 [76] | South Korea | Parallel groups trial | 60 | 72 ± 6 | 100 | Community-dwelling older adults with knee OA | No. of conditions (index condition OA + at least 1 additional condition) | Not reported |
| Ory et al. 2015 [77] | USA | Single group, pre-post | 186 | 75 ± 8 | 84 | Community dwelling older adults | No. of conditions (presence of 2 or more medical conditions) | Not reported |
| Pizarro-Mena et al. 2022 [78] | Chile | Single group pre-post | 45 | 71 ± 8 | 82 | Community dwelling older adults | Defined by authors as multimorbidity (number of conditions) | Not reported |
| Potiaumpai et al, 2024 [79] | USA | RCT | 82 | 61 ± 12 | 35 | Hematopoietic stem cell transplant recipients | No. of conditions (index condition cancer + at least 1 additional condition) | Not reported |
| Rauzi et al. 2024 [80] | USA | Parallel groups | 50 | 69 ± 7 | 36 | Military veterans with multimorbidity | Identified by authors as having multimorbidity | Not reported |
| Reeves et al. 2017 [81] | USA | Pilot RCT | 27 | 72 ± 10 | 59 | Patients with heart failure | No. of conditions (index condition heart failure + at least 1 additional condition) | Not reported |
| Rimmer et al. 2000 [82] | USA | Parallel groups | 35 | 53 ± 8 | 74 | Patients with previous stroke | No. of conditions (index condition previous stroke + at least 1 additional condition) | Not reported |
| Rimmer et al. 2002 [83] | USA | Parallel groups | 44 | 54 | 84 | Community-dwelling adults | Defined by authors as multimorbidity (number of conditions) | Not reported |

| | | | | | | | | |
|---|---|---|---|---|---|---|---|---|
| Rodriguez-Manas et al. 2019 [84] | Belgium, Czech Republic, UK, France, Germany, Italy, Spain | Cluster RCT | 964 | 78 ± 5 | 49 | Patients aged > 70 years with Type 2 diabetes mellitus | No. of conditions (index condition T2DM + at least 1 additional condition) | Not reported |
| Ryan et al. 2022 [85] | Ireland | Uncontrolled pre-post, pilot study | 19 | 62 ± 35 | 74 | Patients with multimorbidity | Defined by authors as multimorbidity (No. of conditions) | Not reported |
| Seino et al. 2017 [86] | Japan | Randomised, controlled crossover trial | 77 | 76 ± 6 | 31 | Community-dwelling older adults | No. of conditions (presence of 2 or more medical conditions) | Not reported |
| Serra-Prat et al. 2017 [87] | Spain | RCT | 172 | 78 ± 5 | 56 | Community-dwelling older adults | No. of conditions (presence of 2 or more medical conditions) | Not reported |
| Servantes et al. 2011 [88] | Brazil | RCT | 45 | 52 ± 9 | 47 | Patients with chronic heart failure | No. of conditions (index condition of CHF + at least 1 additional condition) | Not reported |
| Shiozaki et al. 2023 [89] | Japan | Single group pre-post | 24 | 82 ± 5 | 67 | Frail older patients with multimorbidity | Defined by authors as multimorbidity (number of conditions) | Not reported |
| Soria-Comes et al, 2024 [90] | Spain | Single group, pre-post | 22 | Median age 68 (IQR 64-76) | 32 | Lung cancer survivors | No. of conditions (index condition cancer + at least 1 additional condition) | Cumulative illness rating scale - Geriatric |
| Stevens-Lapsley et al. 2023 [91] | USA | RCT | 150 | 77 ± 9 | 15 | Community-dwelling veterans | Functional comorbidity index | 18-item list of diagnoses |
| Tao et al. 2015 [92] | China | RCT | 113 | 55 ± 11 | 48 | Hemodialysis patients | No. of conditions (Index condition renal failure + at least 1 additional condition) | Not reported |
| Teuwen et al [93] | Netherlands | RCT | 217 | 59 ± 13 | 90 | Adults with rheumatoid arthritis and severe functional limitations | No. of conditions (index condition RA + at least 1 additional condition) | Not reported |
| Tikkanen et al. 2013 [94] | Finland | RCT | 559 | 81 ± 4 | 68 | Community-dwelling older adults | Functional comorbidity index | 13-item list of diagnoses |
| Ullrich et al. 2022 [95] | Germany | RCT | 118 | 82 ± 6 | 76 | Community-dwelling older adults with mild to moderate cognitive impairment | Defined by authors as multimorbidity (number of conditions) | Not reported |
| Van den Helder et al. 2020 [96] | Netherlands | Cluster RCT | 224 | 72 ± 7 | 71 | Community-dwelling older adults | No. of conditions (presence of at least 2 medical conditions) | Not reported |

| | | | | | | | | |
|---|---|---|---|---|---|---|---|---|
| Weiner et al. 2023 [97] | USA | RCT | 99 | 68 ± 8 | 25 | Community-dwelling adults with CKD | No. of conditions (index condition CKD + at least 1 additional condition) | Not reported |
| Wilson et al. 2021 [98] | Australia | Single group, pre-post | 14 | 72 ± 9 | 0 | Patients with prostate cancer on androgen deprivation therapy | Mean number of conditions (Index condition prostate cancer + at least 1 additional condition) | Not reported |
| Winters-Stone et al. 2023 [99] | USA | RCT | 442 | 62 ± 6 | 100 | Cancer survivor patients at risk for falls | Functional comorbidity index | 18-item list of diagnoses |
| Zhou et al. 2021 [100] | Sweden | RCT | 112 | 67 ± 13 | 32 | CKD patients | Davies comorbidity score (Index condition CKD) | 7 domains of active comorbid disease |

*mean ± standard deviation (SD) unless otherwise stated
*CHF* Chronic heart failure, *CKD* Chronic kidney disease, *COPD* chronic obstructive pulmonary disease, *HL* Healthy lifestyle, *ILI* Intensive lifestyle intervention, *OA* Osteoarthritis, *RCT* Randomised controlled trial, *T2DM* Type 2 diabetes mellitus

**Table S3: Exercise intervention details (ordered alphabetically by first author surname)**

| | Resistance exercise programme | | | | | |
|---|---|---|---|---|---|---|
| Study | Intervention duration | Session duration and frequency | Exercise characteristics | Exercise mode(s) | Delivery mode | Other exercise modes included |
| Al-Mhanna et al. 2024 [33] | 12 weeks | 20-60 min x 3 times per week | 5 strength exercises; weeks 1–6, 2 x 15 repetitions with 1 min rest between exercises and rounds, weeks 7–12, 4 x 30 repetitions.<br>*Progression:* increasing weight and RPE (weeks 1–6: RPE 11–13; weeks 7–12: RPE 14–16) | Bodyweight and adjustable dumbbells | Home-based, individual | Aerobic |
| Barker et al. 2018 [34] | 8 weeks | 60 min x 2 times per week | Four upper and three lower limb resistance exercises at 10-12 RM<br>*Progression:* based on target RPE of 12-14 | Free weights | Individual, performed in an outpatient setting, supervised | Aerobic |
| Baptista et al. 2018 [35] | 24 months | 60 min x 3 times per week | 5-8 exercises for the large muscle groups with 1-3 sets of 8-12 repetitions at 50-70% 1RM.<br>*Progression:* based on duration, volume, or intensity every 6 weeks. | Body weight, free weights | Supervised, group-based, delivered in the community | Aerobic, balance, flexibility |
| Bennett et al. 2020 [36] | 3 months | 1-2 times per week | 3 upper- and lower-body exercises for 2 sets of 8-12 repetitions.<br>Progression involved increases in number of exercises, sets, and repetitions. | Resistance bands | Supervised at dialysis centre | Aerobic |
| Bernocchi et al. 2018 [37] | 4 months | 3-7 times per week | Calisthenics and muscle reinforcement exercises using 0.5 kg weights<br>Intensity described as moderate to high level of muscle fatigue | Body weight and free weights | Individual, home-based, initial session supervised, otherwise unsupervised with weekly follow-up by phone | Aerobic |
| Berry et al. 2003 [38] | Short-term intervention: 3 months<br>Long-term intervention: 18 months | 60 min x 3 times per week | Five upper-body strength exercises; 2 sets of 8-10 repetitions<br>*Progression:* not reported | Not reported | Group-based at exercise centre, supervised | Aerobic |
| Blasco-Lafarga et al. 2021 [39] | 6 months | 50 min x 3 times per week | Exercises, volume, and intensity were conditioned to the patients' capacities, classified into high, medium, or low level, according to their dependence status, health, technical capacities, and movement velocity | Elastic bands, small weights, instability cushions, ropes, balls | Supervised & unsupervised | Balance, mobility, co-ordination |

| | | | | | | |
|---|---|---|---|---|---|---|
| Bourne et al. 2017 [40] | 6 weeks | 2 supervised + 3 home-based sessions per week | 10 exercises (including bicep curls, squats, leg extensions, upright row) starting from 60s in week 1, progressing to 3½ min in week 6 | Free weights | Group sessions in a local rehabilitation facility or online PR. Supervised an unsupervised | Aerobic |
| Buford et al. 2012 [41] | 12 months | 40-60 min x 1-3 per week plus home-based activity | Lower-body strength exercises Intensity of RPE 15 to 16 | Ankle weights | Combination of supervised centre- and unsupervised home-based | Aerobic, balance, flexibility |
| Celli et al. 2022 [42] | 12 months | 30 min x 3 times per week | 9 upper- and lower-body resistance exercises. 1-3 sets of 8-12 repetitions at 65-85% of 1RM | Resistance exercise machines | Supervised centre-based (6 months) and independent home-based (6 months) | Aerobic, balance |
| Chang et al. 2017 [43] | 12 weeks | 3 times per week | Seated, open chain lower-body exercises 5 sets of 10 repetitions at 10RM (RPE 13) Intensity progressed by increasing the strength of resistance bands | Resistance bands | Home-based, individual | None |
| Charikiopoulou et al. 2019 [44] | 13 weeks | 2 times per week | 3 sets x 10-15 repetitions for 6-8 exercises of main muscle groups. RPE used to regulate exercise intensity. | Not reported | Individual, centre-based, supervised | Aerobic, respiratory muscle training |
| Chen et al. 2020 [45] | 12 weeks | 10 min x 3 times per week | 2 sets of 10 repetitions Lower-body exercises Intensity progressed by increasing the strength of resistance bands | Resistance bands | Home-based, unsupervised | Aerobic |
| Choi et al. 2024 [46] | 12 weeks | 3 times per week | Whole-body resistance training (6 exercises). Intensity prescribed using RPE (weeks 1-2, 10 to 12; weeks 3-12, 12-14 on the Borg scale) *Progression:* Number of exercises, sets and reps increased with increases in resistance band tension | Bodyweight and resistance bands | Supervised at university gym | None |
| Coca-Martinez et al. 2024 [47] | 12 weeks | 1 supervised session per week + 3 home-based sessions per week | Supervised in-hospital exercise: 10 exercises targeting all major muscle groups with lower-body focus Home-based: whole-body muscular training focused on lower-body using elastic bands | Elastic bands | Supervised, in-hospital & home-based independent exercise | Aerobic |
| Dao et al. 2013 [48] | 12 months | 60 mins x 1-2 times per week | Progressive and high-intensity resistance training | Resistance exercise machines and free weights | Not reported | None |
| Edelmann et al. 2011 [49] | 3 months | 2 times per week | 60-65% of 1RM. 15 repetitions per exercise. 6 exercises targeting major muscle groups. | Free weights and resistance exercise machines. | Individual, centre-based, supervised | Aerobic |

| | | | | | | |
|---|---|---|---|---|---|---|
| Felcar et al. 2018 [50] | 6 months | 2-3 sessions per week totalling 60 sessions | strength training for lower limbs (quadriceps) and upper limbs (biceps and triceps): 3 sets of 8 repetitions at 70% 1RM with load increased weekly. | Not reported | Supervised, group-based | Aerobic |
| Galvao et al. 2014 [51] | 12 months | 2 times per week | Upper- and lower-body resistance training performed for 2-4 sets at 6-12 RM *Progression:* increases in volume and intensity | Not reported | Months 1-6: supervised, group-based; Months 7-12: unsupervised at home | Aerobic |
| Gary et al. 2011 [52] | 12 weeks | 60-90 min x 2-3 times per week | Upper- and lower-body resistance training performed for 2-3 sets of 12-15 repetitions *Progression:* Exercise prescription adjusted at each home visit | Resistance bands | Supervised, home-based | Aerobic |
| Genest et al. 2021 [53] | 6 months | 30 min x 2 times per week | A total of 6 upper- and lower-body exercises with a focus on trunk strength. Exercises performed as many reps as possible in 2 minutes. *Progression:* Weight increased every 3 weeks | Resistance exercise machines | Supervised, group-based | None |
| GilHerrero et al. 2022 [54] | 12 weeks | 50-60 min x 2 times per week | Upper- and lower-body resistance exercise performed for 2-4 sets of 8-12 repetitions at 65-85% 1RM. *Progression:* intensity gradually increased over the intervention | Resistance exercise machines | Two supervised sessions per week plus 1 individualized home-based session | Aerobic |
| Hall et al. 2020 [55] | 12 weeks | 60-90 min x 3 times per week | 1–2 sets of 8–15 repetitions for 8-12 exercises. Moderate to hard intensity. | Body weight, free weight, cables and exercise bands | Supervised, group exercise in community | Aerobic, flexibility, balance |
| Hellberg et al. 2019 [56] | 12 months | 30 min x 3 times per week | 4-6 exercises for upper and lower body performed for 2-3 sets x 10 repetitions. *Progression:* Increasing weight or changing body position | Not specified | Unsupervised, individual exercise at home or at a gym | Aerobic |
| Hetherington et al. 2018 [57] | 24 weeks | 45 min x 2 times per week | Upper- and lower-body resistance exercise performed for 3 sets of 8-12 repetitions at moderate to high intensity (up to approximately 75% 1RM) | Air-pressure driven, computer-integrated machines | Community-based, supervised older-adult-specific exercise clinic | Balance |
| Hinrichs et al. 2016 [58] | 12 weeks | Target number of exercise sessions was 2 or more times per week | Upper- and lower-body RE for 3 sets of 15 repetitions performed at moderate intensity (Perceived level of effort: 5-6 on a 10-point scale) *Progression:* Exercise intensity was regularly adapted throughout the intervention | Body weight and elastic resistance bands | Individual, home-based, unsupervised | Aerobic, balance, flexibility |
| Hjermundrud et al. 2024 [59] | 4 weeks | 60 min x 2 sessions per day (2 strength sessions per week) | Upper- and lower-body strength training. Six exercises per session for 12 repetitions targeting hip extensors, flexors, adductors and abductors; core muscles and upper-arm extensors and flexors | Free weights, resistance bands, strength training machines | Supervised group and individual sessions | Aerobic, balance |

| | | | | | | |
|---|---|---|---|---|---|---|
| Houben et al. 2024 [60] | 20 weeks | 60 min x 2 per week | 4 x 10 repetitions of leg press and leg extension (65-70% 1RM) separated by upper-body exercises *Progression:* training load adjusted based on repeated 1RM testing | Strength training machines | Hospital based, supervised for 20 weeks; followed by continuing home-based exercise up to 1 year | None |
| Jacobs et al. 2014 [61] | 12 weeks | 60 min x 3 times per week | Upper- and lower-body RE for 3 sets of 15 at 60-70% 1RM *Progression:* Intensity progressed every 2 weeks | Free weights, resistance exercise machines, ankle weights | Not reported | Aerobic, flexibility, balance |
| Kaye et al. 2020 [62] | 4 weeks | 90 min x 3 times per week | Full body strength training for 1-2 sets of 12-15 repetitions *Progression:* Weight increased when participants reached 15 repetitions in the second set | Resistance exercise machines, medicine balls. | Individual, supervised, community-based | Aerobic, stretching |
| Kelly et al. 2016 [63] | 7 weeks | 12 sessions in 7 weeks (goal of 2 sessions per week) plus daily home exercise | 1-2 sets of 10 reps at 50-80%1RM *Progression:* increase in intensity | Open and closed chain strengthening exercises | Group-based, supervised & individual, home-based | Aerobic, stretching |
| Kitzman et al. 2021 [64] | 12 weeks | 60 min x 3 sessions per week | Strengthening exercises of the lower extremities supplemented by general resistance exercises; 1 set x 10 repetitions at RPE <12-15/16 | Resistance bands, free weights | Individual, supervised | Balance, aerobic, mobility |
| Krumov et al. 2022 [65] | 6 months | Not reported | Progressive strengthening exercises (knee flexion, mini-squats, step-up, upper limb weights) | Not reported | Group-based, supervised | Balance, gait training |
| Lauze et al. 2017 [66] | 12 weeks | 45 min x 2 times per week | 8 resistance exercises performed at "Light to moderate" intensity | Body weight, tech equipment included computer with software and motion capture device | Individual, home-based, hybrid; 6 of 24 sessions were supervised | Aerobic, balance |
| Layne et al. 2009 [67] | 12 weeks | 60 min x 2 times per week | 2 sets x 8 repetitions. Target RPE 6-8 "hard" to "very hard" | resistance exercise equipment such as free weights, cuff weights and dumbbells | Group-based, supervised at local fitness center | Balance, flexibility |
| Lee et al. 2022 [68] | 12 months | 30 min x 16 sessions over 1 year | Not reported | At instructor's discretion (eg, resistance band/weight, exercise ball, free-weight postures; one-legged stand, tandem walk) | Supervised, group-based | Stretching, flexibility |
| Legault et al. 2025 [69] | 2-8 weeks | 55-60 min x 3 times per week | Seven functional movements (e.g., squats) targeting the whole-body for 30-60s with 30-60s rest between exercises *Progression:* RPE based | Body weight, resistance bands | SPP group: all sessions supervised SSPP group: 1 x supervised, 2 x home-based per week | Aerobic |

| | | | | | | |
|---|---|---|---|---|---|---|
| Mangani et al. 2006 [70] | 18 months | 60 min x 3 times per week | 2 sets x 12 repetitions of 9 exercises<br><br>Intensity not specified | Dumbbells and ankle weights | Individual, facility-based (first 3 months). Home-based (second 15 months). Hybrid; 3 months supervised, 15 months unsupervised | None |
| Martin-Alemany et al. 2022 [71] | 6 months | Not reported | 4 sets x 20 repetitions of 4 exercises (leg extension, arm extension, straight leg extension, seated marching).<br>*Progression:* increasing ankle weights or strength of resistance band | Resistance bands, ankle weights | Not reported/unclear | Aerobic |
| Merchant et al. 2021 [72] | 3 months | 60 min x 1-2 times per week | Resistance exercises performed in a circuit | Body weight | Supervised, group-based, at community center, | Aerobic, balance |
| Mulrow et al. 1994 [73] | 4 months | 30-45 min x 3 times per week. | 69% of sessions included strength exercise performed up to 15 repetitions | Elastic bands and free weights | Individual, at the nursing home, supervised | Aerobic, balance, coordination training |
| Nemoto et al. 2024 [74] | 48 weeks | 60 min, once every 2 weeks (10-20 min strength training) | Whole-body strength exercises – upper- and lower-body focused performed at moderate intensity<br>Participants also instructed to perform regular exercises at home | Strength training machines | Individual, supervised hospital based, home-based | Aerobic, balance, flexibility, coordination |
| O'Brien et al. 2021 [75] | 6 months | 90 min x 3 times per week (30 min RE per session) | 60-70% 1RM. 1 set with 10-12 repetitions 8-10 exercises for major muscle groups | Resistance exercise equipment such as resistance exercise machines | Both group and individual exercise, at community center, Hybrid, weekly supervised sessions | Aerobic, balance, flexibility |
| Oh et al. 2021 [76] | 5 months | 60-80 min x 2-3 times per week | 12 different movements<br>low-resistance bands were used initially. Intensity gradually increased but levels not specified. | Elastic bands | Self-directed home-based. Hybrid, monthly supervised sessions | None |
| Ory et al. 2015 [77] | 10 weeks | 90 min x 2 times per week | 15 minutes of strength exercise | Body weight | Group-based, at community center, supervised | Aerobic, balance, flexibility |
| Pizarro-Mena et al. 2022 [78] | 12 weeks | 60 min x 3 times per week | 1-4 sets of 8-15 reps at 60-80% 1RM | Dumbbells, ankle weights, elastic bands | Face to face, group, supervised | Aerobic, flexibility, balance |
| Potiaumpai et al, 2024 [79] | 2-24 weeks | 30-45 mins x 5 times per week | 4-6 upper- and lower-body strength exercises.<br>*Progression:* increasing or decreasing sets, repetitions, weight, and/or the number of exercises prescribed | Body weight and resistance bands | Home-based | Aerobic |
| Rauzi et al. 2024 [80] | 12 weeks | 60 min x 2-3 times per week | Strengthening exercises performed at intensity of 8RM<br>*Progression:* Intensity progressed when participants could perform 8 reps without signs of technical failure | Weights, resistance bands | Combination of individual and group & supervised and remote delivery | None |

| | | | | | | |
|---|---|---|---|---|---|---|
| Reeves et al. 2017 [81] | 12 weeks | 60 min x 3 times per week | Functional strength training focused on lower body (chair rises, step-ups) Self-perceived intensity at RPE 15-16 | Body weight | Supervised, group-based at rehabilitation facility, Home-based on in-between days, hybrid | Balance, mobility, aerobic |
| Rimmer et al. 2000 [82] | 12 weeks | 60 min x 3 times per week | Combination of 8 upper- and lower-body exercises performed 1 set of 15-20 repetitions at 70% of 10RM | Resistance exercise machines | Group-based, at university fitness center, supervised | Aerobic, flexibility |
| Rimmer et al. 2002 [83] | 12 weeks | 60 min x 3 times per week | 1 set of 15-20 repetitions performed at 70% of 10RM | Resistance exercise machines | Group-based, at community center, supervised | Aerobic, flexibility |
| Rodriguez-Manas et al. 2019 [84] | 16 weeks | 45 min x 2 times per week | 40-80% of 1RM, 2-3 sets of 8-10 repetitions | Resistance exercise machines. Two exercises performed: leg press and bilateral knee extension | Individual, centre-based, supervised | None |
| Ryan et al. 2022 [85] | 6 weeks | 45 min x 1 time per week | A range of strength exercises self-selected by the participants (RPE 5-8). Exercise volume increased overtime | Body weight, hand weights and lower limb exercise machines | Group-based, at community leisure centre, supervised | None |
| Seino et al. 2017 [86] | 3 months | 60 min x 2 times per week | 2 sets x 20 repetitions (6 upper- and lower-body exercises) at self-perceived intensity of “somewhat hard”. Sets are repetitions were increased progressively. | Body weight and elastic bands | Group-based, at local community center, supervised | None |
| Serra-Prat et al. 2017 [87] | 12 months | 20-25 min x 4 times per week | 10 strengthening exercises performed for 10-15 repetitions per minute, 30 s rest in between | Free weights, resistance exercise equipment and body weight | Individual, home-based. Unsupervised (1 introduction session) | Aerobic, balance, coordination |
| Servantes et al. 2011 [88] | 3 months | 20 min x 3-4 times per week | Three exercises for upper limbs and four exercises for lower limbs. 30-40% of 1RM, 1 set of 12-16; progression based on increasing repetitions or weight | Free weights | Individual, home-based. Hybrid (first three sessions supervised) | Aerobic |
| Shiozaki et al. 2023 [89] | 4 weeks | Resistance training: 60 min x 3-6 times per week | Leg press, leg extension, hip abduction. 2-4 sets of 15 repetitions, 40-60% 1RM | RE machines | Individual inpatient exercise, supervised | Aerobic, flexibility |
| Soria-Comes et al, 2024 [90] | 3 months | 60 min x twice weekly | 3 x 3 sets of upper- and lower-limb exercises<br>*Progression:* Increase in perceived effort from 4-5/10 to 9-10/10 | Not reported | Supervised community exercise delivered at council care centres | Aerobic, balance |
| Stevens-Lapsley et al. 2023 [91] | 1 month | 3 times per week; 12 sessions in total | Progressive high-intensity resistance exercise: 3 sets at 8RM<br>Standard PT: 10 repetitions, no weights | Lower extremity muscle exercises performed with resistance exercise machines/equipment | Individual, home-based, supervised | Aerobic |
| Tao et al. 2015 [92] | 12 weeks | 20 min x 1-2 times per week | Not reported | Body weight and elastic bands | Group-based and later individual, hybrid | Flexibility |
| Teuwen et al [93] | ≥52 weeks | 2 x per week for 12 weeks followed by 1 x per week for 40 weeks | Muscle strengthening exercises<br>*Progression:* intensity was increased 5%-10% increase per week | Bodyweight, machines | Supervised | Aerobic, balance, flexibility |

| | | | | | | |
|---|---|---|---|---|---|---|
| Tikkanen et al. 2013 [94] | 24 months | 75 min x 1 time per week | 60-85% of 1RM, 8-12 repetitions x 2-3 sets | Resistance exercise machines and resistance exercise equipment | Individual, gym-based, supervised | Balance |
| Ullrich et al. 2022 [95] | 12 weeks | Daily (time per session not specified) | Tiptoe stance, sit-to-stand transfers/chair-rise, stair riseIntensity not reported | Body weight | Individual, home-based, hybrid | Aerobic, balance |
| Van den Helder et al. 2020 [96] | 6 months | 45 min x 2-3 times per week | Functional exercises focusing on daily activities. Moderate to vigorous intensity | Body weight and objects at home | Individual, home-based, hybrid | Aerobic, balance, flexibility |
| Weiner et al. 2023 [97] | 12 months | Resistance exercise: 10 minutes x 3 times per week first 6 months | Upper and lower extremity resistance training exercises. Intensity 15-16 RPE | Body weight, ankle weights and hand weights | Group-based and individual, at exercise facility and at home Hybrid; first 6 months supervised, last 6 months 1 out 3 sessions performed at home | Aerobic |
| Wilson et al. 2021 [98] | 12 weeks | 60 min x 3 times per week | 6-12 repetitions x 1-4 sets targeting the major muscle groups of upper- and lower-body | Resistance exercise machines | Individual, gym-based, supervised | Aerobic |
| Winters-Stone et al. 2023 [99] | 6 months | 60 minutes x 2 times per week | Progressive lower-body strength training; 1-3 sets of 8-10 exercises, 8-12 repetitions. | Weighted vests and steps | Group-based, delivered at exercise facility, supervised | None |
| Zhou et al. 2021 [100] | 12 months | 30 min x 3 times per week | 4-6 exercises for upper and lower body performed for 2-3 sets x 10 repetitions. *Progression:* Increasing weight or changing body position | Not specified | Unsupervised, individual exercise at home or at a gym | Aerobic |

*1RM* 1-repetition maximum, *8RM* 8-repetition maximum, *10RM* 10-repetition maximum, *RM* repetition maximum, *RPE* Rating of perceived exertion

**Table S4: Study outcomes (ordered alphabetically by first author surname)**

| Study, year | Outcomes | Main findings | Adverse events | Intervention Adherence & Fidelity |
|---|---|---|---|---|
| Al-Mhanna et al. 2024 [33] | 30s-CST, TUG | Significant pre-post improvements in 30s-CST (18-56%) and TUG (24-29%). | No injuries or adverse effects were reported | 91.7% session attendance |
| Barker et al. 2018 [34] | 6MWT | Both the multimorbidity specific rehabilitation group (22 metres; 95% CI -16 to 60) and the disease specific rehabilitation group (22 metres; 95% CI − 69 to 11) reported an improvement in 6MWT. | No adverse events reported | 63% completed the intervention (defined as 12 of 16 sessions) |
| Baptista et al. 2018 [35] | 30s-CST, TUG, 6MWT | Significant within-group improvements (moderate – large effect sizes) in all outcomes in both exercise groups (Exercise alone & exercise + oral hypertensive drugs). | No serious adverse events reported except for soreness | N/A |
| Bennett et al. 2020 [36] | 30s-CST, TUG | Significant improvement in TUG performance (intervention, −1.7 ± 2.9; control, −0.8 ± 1.6; P = 0.04). No difference in chair stand test. | No serious adverse events caused by the exercise program were reported. A total of 4 events were reported in the exercise group with 2 of these related to the exercise<br>Intervention (minor abdominal Discomfort, dizziness) | 77% completed >50% of exercise sessions |
| Bernocchi et al. 2018 [37] | 6MWT | The intervention group improved 6MWD by 60 m (95% CI 22.2-97.8), compared with no improvement in control group. There was a significant difference in Δ6MWT between the groups. | No adverse events reported | 93% patients performed the prescribed exercises (19% performed 2.3 sessions per week, 65% performed 4 sessions per week, 16% performed 6 sessions per week) |
| Berry et al. 2003 [38] | 6MWT | Participants in the long-term intervention group (18-month) improved 6MWT by 6% compared with short-term intervention group | N/A | Short-term intervention group attended 89% of all possible sessions in the first 3 months (long-term group attended 88% of all sessions in this period). At 18-months the long-term group had attended 52.3% of all sessions. |
| Blasco-Lafarga et al. 2021 [39] | Gait speed, HGS, TUG, 30s-CST | No improvement in handgrip strength, TUG and gait speed. Significant improvement in 30s CST performance after the supervised training period. | N/A | N/A |
| Bourne et al. 2017 [40] | 6MWT | Improvements in 6MWT performance in face-to-face and online delivery groups | Back pain n = 2, Muscular skeletal chest pain n = 1, Inguinal pain n = 1, Common cold n = 1 | 72% of the two face-to-face sessions attended, 62% of suggested 5 sessions recorded as accessed online |
| Buford et al. 2012 [41] | SPPB | Improvements in SPPB score of ACEi users and of those using other anti-hypertensive drugs. | N/A | Median session attendance was 72% for ACEi users, 68% for other users & 70% for non-users |
| Celli et al. 2022 [42] | 1RM, gait speed, lean body mass (DXA) | Strength and gait speed improved more in the intensive lifestyle intervention group compared with the healthy lifestyle group. | 30 events of hypoglycaemia in the intensive lifestyle intervention group and 20 hypoglycaemia events in the healthy lifestyle group | Median attendance was 87% for diet therapy sessions & 91% for exercise therapy sessions |

| | | | | |
|---|---|---|---|---|
| Chang et al. 2017 [43] | TUG, 6MWT | Significant improvements in TUG and 6MWT for baseline vs. week 12. | No report of intervention-related adverse effects | Average exercise adherence rate was 73% |
| Charikiopoulou et al. 2019 [44] | 6MWT | Multimorbidity group improved 6MWT from baseline to follow up. | No adverse events reported | N/A |
| Chen et al. 2020 [45] | 30s-CST, TUG | Both dynamic and isometric resistance groups improved TUG performance compared with baseline. The improvement was greater in the dynamic group. For 30s-CST, both groups improved compared with baseline, with greater effects in the dynamic resistance group. | No adverse events reported | Adherence was 83% in the dynamic resistance group and 88% in the isometric resistance group. |
| Choi et al. 2024 [46] | HGS, 30s-CST, TUG, 6MWT, SMM via BIA | Descriptive analysis only presented. Improvements in HGS, CST, 6MWT and TUG. Skeletal muscle mass decreased. | No adverse events occurred during the intervention | 88.9% session attendance |
| Coca-Martinez et al. 2024 [47] | 6MWT | Significant improvement in the intervention group vs. control (mean between group difference 71m) | N/A | 83% adherence to in-hospital supervised exercise; 90% adherence for home-based exercise |
| Dao et al. 2013 [48] | Lean mass (DXA) | Both once weekly RE and twice weekly RE groups showed reductions in total lean mass post intervention, although no effect sizes or statistical testing are reported. | N/A | N/A |
| Edelmann et al. 2011 [49] | 6MWT | Exercise group improved 6MWT by 24 m (95% CI 10 to 38), which was not significantly different from the control group (17 m, 95% CI -3 to 38) | No serious adverse events; 11 participants (25%) in the exercise group experienced events without clinical relevance (palpitations, dyspnea, musculoskeletal discomfort) | 34% of participants participated in >90% of exercise sessions, 52% in 70-90% of exercise sessions, and 14% in <70% of exercise sessions. None lost to follow-up |
| Felcar et al. 2018 [50] | 6MWT,1RM strength (biceps, triceps, quadriceps), FFM (via BIA) | Improvements in 6MWT and all 1RM values. No change in FFM. | N/A | N/A |
| Galvao et al. 2014 [51] | 400-m walk, 5-CST, 1RM (chest press and leg extension), appendicular muscle mass (DXA) | The exercise group showed a significant improvement in 400m walk, CST and 1RM strength. No changes in appendicular muscle mass at 12 months. | One participant with preexisting back pain elected to cease the exercise programme, as did one patient with a preexisting knee injury. | The exercise group completed a mean of 40 plus or minus 12 of the 52 exercise sessions (77% attendance) |
| Gary et al. 2011 [52] | HGS, 6MWT, upper- (forearm flexion) and lower-body isometric muscle strength (knee extension) | Between group analysis showed significantly greater improvements for the exercise group compared to control for upper-body isometric strength and right leg knee extension. No significant differences between groups for changes in 6MWT or HGS (HGS improved in the Ex group). | N/A | Adherence was 83% and 99% for the walking sessions and resistance exercises, respectively |
| Genest et al. 2021 [53] | HGS, SPPB, 5-CST, Gait speed, TUG, 6MWT, Skeletal muscle index (BIA) | In the resistance training group, there was no change in HGS, 5-CST, SPPB, TUG or skeletal muscle index. There were significant improvements in gait speed and 6MWT | No adverse events reported | Exercise adherence was 71% in the resistance training group |
| GilHerrero et al. 2022 [54] | 1RM (chest, leg press), skeletal muscle mass (BIA) | Significant improvements in chest and leg strength and skeletal muscle mass. | N/A | Median adherence to the supervised exercise program was 80% with a range of 58% to 100% |

| | | | | |
|---|---|---|---|---|
| Hall et al. 2020 [55] | 6MWT, gait speed, 30s-CST, TUG | Moderate beneficial effect for exercise group for 6MWT. Small to large beneficial effects for the other outcomes for the exercise group compared to control. | N/A | 89% of participants completed the intervention, attending 82% of the sessions |
| Hellberg et al. 2019 [56] | 6MWT, 30s-CST, HGS, Isometric quadriceps strength | Significant improvements post-intervention for 6MWT, 30s-STS and quadriceps strength. No change in HGS. There were no treatment differences comparing strength group vs. balance group. | No exercise related side-effects or harm were reported. | In the strength group, 84% of participants reported training at 4 months, 70% at 8 months, and 62% at 12 months. |
| Hetherington et al. 2018 [57] | SPPB | Significant improvement (12%) in SPPB post intervention | N/A | N/A |
| Hinrichs et al. 2016 [58] | 5-CST, TUG, HGS | Experimental group did not significantly improve in any measure vs. control group | 45% of intervention participants and 50% of control participants reported 1 or more adverse events. A total of 151 AEs were reported, out of which 21 (14%) were classified as serious. Experimental intervention was judged to have caused two nonserious AEs | Median intervention time was 84 days. Mean number of exercise sessions per week was 3.7 |
| Hjermundrud et al. 2024 [59] | 10-m walk test | Significant improvement in 10-m walk speed | N/A | 75-80% compliance across the two exercise groups |
| Houben et al. 2024 [60] | Lean mass via DXA, Muscle mass via CT, 1RM (leg press, leg extension) | Muscle strength significantly increased in EX (leg press 4 ± 11%, leg extension 5 ± 16%) and decreased in CON (leg press −10 ± 9%, leg extension −11 ± 13%). Quadriceps muscle CSA decreased in the total population (−2.2 ± 2.9 $cm^2$) with no significant differences between groups. No significant differences between groups over time were observed for ALM. | N/A | N/A |
| Jacobs et al. 2014 [61] | Lean mass (MRI) | Thigh lean tissue area did not change over the intervention period. Over 9-month follow-up, participants in the traditional RE group lost a significant amount of lean tissue, while participants in the eccentric RE group did not. | N/A | Traditional RE group: <50% attendance n = 2, 50-80% n = 2, >80% n = 34<br>Eccentric RE group: <50% attendance n = 4, 50-80% n = 5, > 80% n = 30 |
| Kaye et al. 2020 [62] | TUG, gait speed, 6MWT, lean mass (via air-displacement plethysmography) | Following the intervention there were significant improvements in 6MWT (+58.5 ft), gait speed (-0.36s), TUG (-0.5s) but no change in lean mass. | No adverse events reported | Successful competition, defined as adhering to >70% of sessions was achieved by 80% of participants. Mean sessions completed per patient was 82%. |
| Kelly et al. 2016 [63] | 6MWT, TUG | Both high velocity and low velocity RE groups improved 6WMT and TUG at post-intervention compared with baseline. There were no differences between groups for either outcome. | Two minor adverse events occurred during the study which required a decrease in the intensity of resistance to continue exercise (hamstring soreness & recurrence of previously episodic sciatica) | Participants completed 1.9 sessions per week. |
| Kitzman et al. 2021 [64] | SPPB, gait-speed, 5-CST, 6MWT, HGS | The intervention group had significantly greater improvement in SPPB compared to the control group. Gait speed, 6MWT and 5-CST also improved. There was no change in HGS. | Chest pain, hypertension, dizziness, hyperglycaemia, hypoglycaemia were more common in the intervention group. | Adherence to the sessions was 67% |
| Krumov et al. 2022 [65] | 6MWT | 6MWT performance significantly improved in both group. | N/A | N/A |

| | | | | |
|---|---|---|---|---|
| Lauze et al. 2017 [66] | SPPB, 5-CST, gait speed, TUG, HGS, Muscle mass (BIA) | Significant improvements in TUG, SPPB and gait speed in the exercise group compared to controls. No significant changes in any other outcomes. | Two adverse events (falls) occurred during the intervention which did not lead to any serious injury | Exercise group completed 89% of prescribed exercise sessions. |
| Layne et al. 2009 [67] | 30s-CST, HGS, TUG | Significant improvement in 30s-STS compared to control (32.2% increase in the intervention group). Only within-group (RE group only) improvements for HGS and TUG (+11.6%). | No adverse events reported | Mean adherence to the intervention was 82% |
| Lee et al. 2022 [68] | HGS, gait speed | Intervention improved gait speed in the MIND (mobility impairment, no disability). No change in HGS. | N/A | N/A |
| Legault et al. 2025 [69] | 30s-CST | The minimum clinically important difference in 30s-CST was reached by 30% of participants | No serious adverse events related to the exercise intervention. Mild adverse events (musculoskeletal pain or discomfort) occurred in 52% of the participants. | SPP group: 80 ± 15% sessions attended<br>SSPP group: 73 ± 30% sessions attended |
| Mangani et al. 2006 [70] | 6MWT | Strength training group did not improve 6MWT | N/A | 64% attended 70% or more of the sessions |
| Martin-Alemany et al. 2022 [71] | 6MWT, Gait speed, 5-CST, TUG, HGS, SPPB, thigh muscle area (via CT) | Gait speed, 6MWT and HGS significantly improved in the nutrition + exercise group. There was no improvement in 5-CST, TUG or SPPB and no change in thigh muscle area. | N/A | N/A |
| Merchant et al. 2021 [72] | HGS, SPPB, gait speed | Exercise group significantly improved gait speed and SPPB. No change in HGS. | N/A | Participants attended an average of 13 sessions over 3 months |
| Mulrow et al. 1994 [73] | HGS, Upper- and lower-body isometric strength | No significant improvements in the exercise group | None from exercise sessions | 89% of exercise sessions were completed; 13 participants in the exercise group missed more than 3 weeks of scheduled sessions due to acute illness and hospitalisations. |
| Nemoto et al. 2024 [74] | HGS, TUG, gait speed | Improvement in TUG. No change in HGS and gait speed | No adverse events reported | Mean attendance was 93.6%. Home exercise mean adherence was 68.5%. |
| O'Brien et al. 2021 [75] | HGS | No significant improvement in HGS | No serious adverse events from the intervention reported | Median attendance was 72% |
| Oh et al. 2021 [76] | 5-CST, TUG, gait speed, HGS, knee extensor strength, skeletal muscle mass (method not reported) | Compared to the control group, the intervention group significantly improved 5-STS by 36.4%, TUG by 4.5%, gait speed by 28.3% and knee extensor strength by 4.1%. There were no changes in HGS or skeletal muscle mass. | N/A | Adherence to the exercise programme described as "low" (satisfactory adherence defined as ≥80%) |
| Ory et al. 2015 [77] | TUG | 11% improvement in TUG time | N/A | 60% of participants attended 70% or more of the classes |
| Pizarro-Mena et al. 2022 [78] | SPPB, TUG, 30s-CST, gait speed, HGS | Significant improvements in SPPB, TUG and 30s-STS following the intervention period. No change in HGS or gait speed. | N/A | Average attendance was 84%. |
| Potiaumpai et al, 2024 [79] | 6MWT, SPPB, HGS, TUG, 30s-CST | Significant improvements in 6MWT, SPPB and 30s-CST in the exercise group compared to usual care. No changes in other outcomes. | No adverse events reported | Mean adherence was 92% |
| Rauzi et al. 2024 [80] | 30s-CST | No improvement in 30s-CST performance. | 32% of participants reported one adverse event, 38% reported ≥ 2 events. Falls were the most common adverse event. | 78% of participants adhered to all sessions |
| Reeves et al. 2017 [81] | SPPB, 6MWT | Intervention improved SPPB by 1.1 units and 6MWT performance by 23m. | 1 adverse event possibly related to intervention (myocardial infarction) | Participants who completed the intervention attended 92% of scheduled sessions |

| | | | | |
|---|---|---|---|---|
| Rimmer et al. 2000 [82] | HGS, 10RM (leg press and bench press) | Significant increases in leg press and bench press. No improvement in HGS. | There were 3 adverse events, with one occurring during the exercise programme (dizziness) | Participants attended 93% of exercise sessions |
| Rimmer et al. 2002 [83] | HGS, 10RM (leg press and bench press) | Significant increases in leg press and bench press. No improvement in HGS. | No adverse events related to the exercise were reported | Participants attended 87% of exercise sessions |
| Rodriguez-Manas et al. 2019 [84] | SPPB | The intervention group significantly improved SPPB by 0.85 points compared with the usual care group | Adverse events occurred in 22% of the intervention group. No serious adverse events were related to the exercise intervention | 82% of participants in the intervention group reached the adherence criteria (5 out of 7 nutritional and educational sessions, 23 out of 32 resistance exercise sessions) |
| Ryan et al. 2022 [85] | HGS, 6MWT | Small improvements in HGS and 6MWT reported but no formal analysis conducted. | No adverse events related directly to the programme reported | Attendance was at least 60% at five or more sessions and 75% of participants completed the programme. |
| Seino et al. 2017 [86] | HGS, gait speed, TUG | Intervention group significantly improved TUG by -0.25s. No significant improvements in other outcomes. | No adverse events or injuries reported | The average total adherence rate was 90.4%. |
| Serra-Prat et al. 2017 [87] | HGS, gait speed, TUG | No effect on measures HGS, gait speed or TUG. | No adverse events reported | In the intervention group 47.5% of completers adhered “well”; 24% of participants in the intervention group dropped out by follow-up. |
| Servantes et al. 2011 [88] | Isometric knee strength | Knee strength (flexors and extensors) significantly improved in the combined group compared to aerobic exercise alone | No adverse events related to the training reported | Average adherence in the strength/aerobic group was 100% |
| Shiozaki et al. 2023 [89] | HGS, isometric leg strength, 6MWT, 10-m walk test | Participants significantly improved all muscle strength and walking parameters. | No adverse events requiring medical treatment occurred and there were no exacerbations of comorbidities. | Average of days of exercise was 26.5 days. Average total exercise time was 48.9 hours. |
| Soria-Comes et al, 2024 [90] | 30s-CST, HGS, 6MWT, SPPB, Gait speed, Lean mass via BIA | Significant post-intervention improvements in 30s-CST, gait speed, 6MWT and SPPB. No change in HGS and lean mass. | No adverse events reported | Mean attendance to exercise sessions was 70% |
| Stevens-Lapsley et al. 2023 [91] | Gait speed, TUG, SPPB, HGS, knee extensor strength | No statistically significant difference in any outcome between group receiving high-intensity physical therapy and those receiving standard physical therapy | During the intervention period (0-30 days) there were 30 falls in total; 10 falls required medical attention. 22 ED visits and 26 hospitalisations. Unclear if any related to the exercise component | Participants receiving high-intensity PT had a mean of 10.4 home visits. Strength training performed in >80% of prescribed sessions. |
| Tao et al. 2015 [92] | Gait speed, 10-STS | The intervention group significantly improved normal gait speed. Change in 10-STS was not significant between groups although the increase was greater in the intervention group compared to control. | N/A | N/A |
| Teuwen et al [93] | 6WMT | Significant improvement in 6MWT performance (Mean between group difference in change scores 56m) | No serious adverse events reported. One adverse event in the intervention group (dizziness and nausea) | Average 39 sessions attended |
| Tikkanen et al. 2013 [94] | 5-CST | The intervention improved CST in physically active women. There was no improvement in inactive women, inactive men, or active men. | N/A | N/A |
| Ullrich et al. 2022 [95] | SPPB, TUG | The intervention group significantly improved SPPB by 1.9 units (compared to control) and TUG by 5.8 seconds after the 12-week intervention period | Adverse events were associated with preexisting conditions and not directly or indirectly related to the intervention. | Mean training adherence was 60% |

| | | | | |
|---|---|---|---|---|
| Van den Helder et al. 2020 [96] | SPPB, TUG, 6MWT, Gait speed, HGS, skeletal muscle mass (DXA) | No significant effects for SPPB, TUG, 6MWT, HGS or gait speed for exercise vs. control. Exercise + nutrition group improved HGS and gait speed vs. control. No change in skeletal muscle mass. | Five serious adverse events were reported during the intervention period and two during the follow-up, but without relation to the study. | Adherence was 75% in the exercise group and 49% in the exercise + protein group. |
| Weiner et al. 2023 [97] | 6MWT, TUG, SPPB | 6MWT and TUG time was significantly improved in the exercise group compared to control at follow-up (12-months). No difference between groups in SPPB score. | 182 adverse events occurred (40% were deemed as serious and 44% expected due to comorbidities). No serious adverse events were related to the exercise training. | Exercise group completed 59.5% of prescribed exercise sessions. Adherence was lower in the 6-month maintenance phase (48.9%) |
| Wilson et al. 2021 [98] | Total lean mass (DXA), 1RM (leg press and chest press) | No change in lean mass. Significant increase in leg press (24.7%) and chest press (19.8%) | No adverse events occurred during the exercise sessions. One participant experienced an infected leg wound caused by resistance band exercises. | Participants attended 89% of supervised exercise sessions. 11 of 14 patients completed the study. |
| Winters-Stone et al. 2023 [99] | 1RM (leg press) | Strength training group significantly increased their maximal leg strength by 14 kg compared with stretching group after the 6-month intervention period. | 11 adverse events related or possibly related to exercise training (6 mild, 5 moderate, 0 severe). AEs were mainly from the strength training group (n = 9). AE's were pain related exacerbations of existing knee or back pain or new-onset pain. | The strength training group completed 73.3 ± 20.9% of prescribed sessions, with 67% in this group continuing with 1 session per week after the intervention period (months 7-12) |
| Zhou et al. 2021 [100] | Lean mass (DXA) | No change in any lean mass parameter (arms, legs, trunk or whole-body) | No adverse events reported | N/A |

*1RM* 1-repetition maximum, *6MWT* 6-minute walk test, *10RM* 10-repetition maximum, *10-STS* 10-repetition sit-to-stand, *30s-STS* 30-s sit to stand test, *BIA* bioelectrical impedance, *CST* 5-repetition chair stand test, *DXA* Dual-energy X-ray absorptiometry, *FFM* fat free mass, *HGS* Handgrip strength, *MRI* Magnetic Resonance Imaging, *N/A* not reported, *SPPB* Short Physical Performance Battery, *TUG* Timed up-and-go

**Table S5. Preferred Reporting Items for Systematic reviews and Meta-Analyses extension for Scoping Reviews (PRISMA-ScR) Checklist**

| SECTION | ITEM | PRISMA-ScR CHECKLIST ITEM | REPORTED ON PAGE # |
|---|---|---|---|
| **TITLE** | | | |
| Title | 1 | Identify the report as a scoping review. | 1 |
| **ABSTRACT** | | | |
| Structured summary | 2 | Provide a structured summary that includes (as applicable): background, objectives, eligibility criteria, sources of evidence, charting methods, results, and conclusions that relate to the review questions and objectives. | 2 |
| **INTRODUCTION** | | | |
| Rationale | 3 | Describe the rationale for the review in the context of what is already known. Explain why the review questions/objectives lend themselves to a scoping review approach. | 3-4 |
| Objectives | 4 | Provide an explicit statement of the questions and objectives being addressed with reference to their key elements (e.g., population or participants, concepts, and context) or other relevant key elements used to conceptualize the review questions and/or objectives. | 4 |
| **METHODS** | | | |
| Protocol and registration | 5 | Indicate whether a review protocol exists; state if and where it can be accessed (e.g., a Web address); and if available, provide registration information, including the registration number. | 5 |
| Eligibility criteria | 6 | Specify characteristics of the sources of evidence used as eligibility criteria (e.g., years considered, language, and publication status), and provide a rationale. | 5 |
| Information sources* | 7 | Describe all information sources in the search (e.g., databases with dates of coverage and contact with authors to identify additional sources), as well as the date the most recent search was executed. | 6 and Supporting material |
| Search | 8 | Present the full electronic search strategy for at least 1 database, including any limits used, such that it could be repeated. | Supporting material Table S1 |
| Selection of sources of evidence† | 9 | State the process for selecting sources of evidence (i.e., screening and eligibility) included in the scoping review. | 5-6 |
| Data charting process‡ | 10 | Describe the methods of charting data from the included sources of evidence (e.g., calibrated forms or forms that have been tested by the team before their use, and whether data charting was done independently or in duplicate) and any processes for obtaining and confirming data from investigators. | 6 |
| Data items | 11 | List and define all variables for which data were sought and any assumptions and simplifications made. | 6 |
| Critical appraisal of individual sources of evidence§ | 12 | If done, provide a rationale for conducting a critical appraisal of included sources of evidence; describe the methods used and how this information was used in any data synthesis (if appropriate). | N/A |
| Synthesis of results | 13 | Describe the methods of handling and summarizing the data that were charted. | 6 |
| **RESULTS** | | | |
| Selection of sources of evidence | 14 | Give numbers of sources of evidence screened, assessed for eligibility, and included in the review, with reasons | 7 |

| SECTION | ITEM | PRISMA-ScR CHECKLIST ITEM | REPORTED ON PAGE # |
|---|---|---|---|
| | | for exclusions at each stage, ideally using a flow diagram. | |
| Characteristics of sources of evidence | 15 | For each source of evidence, present characteristics for which data were charted and provide the citations. | 7 |
| Critical appraisal within sources of evidence | 16 | If done, present data on critical appraisal of included sources of evidence (see item 12). | N/A |
| Results of individual sources of evidence | 17 | For each included source of evidence, present the relevant data that were charted that relate to the review questions and objectives. | 7-12 |
| Synthesis of results | 18 | Summarize and/or present the charting results as they relate to the review questions and objectives. | 7-12 |
| **DISCUSSION** | | | |
| Summary of evidence | 19 | Summarize the main results (including an overview of concepts, themes, and types of evidence available), link to the review questions and objectives, and consider the relevance to key groups. | 12-16 |
| Limitations | 20 | Discuss the limitations of the scoping review process. | 16 |
| Conclusions | 21 | Provide a general interpretation of the results with respect to the review questions and objectives, as well as potential implications and/or next steps. | 16 |
| **FUNDING** | | | |
| Funding | 22 | Describe sources of funding for the included sources of evidence, as well as sources of funding for the scoping review. Describe the role of the funders of the scoping review. | 17 |

JBI = Joanna Briggs Institute; PRISMA-ScR = Preferred Reporting Items for Systematic reviews and Meta-Analyses extension for Scoping Reviews.
* Where *sources of evidence* (see second footnote) are compiled from, such as bibliographic databases, social media platforms, and Web sites.
† A more inclusive/heterogeneous term used to account for the different types of evidence or data sources (e.g., quantitative and/or qualitative research, expert opinion, and policy documents) that may be eligible in a scoping review as opposed to only studies. This is not to be confused with *information sources* (see first footnote).
‡ The frameworks by Arksey and O'Malley (6) and Levac and colleagues (7) and the JBI guidance (4, 5) refer to the process of data extraction in a scoping review as data charting.
§ The process of systematically examining research evidence to assess its validity, results, and relevance before using it to inform a decision. This term is used for items 12 and 19 instead of "risk of bias" (which is more applicable to systematic reviews of interventions) to include and acknowledge the various sources of evidence that may be used in a scoping review (e.g., quantitative and/or qualitative research, expert opinion, and policy document).

*From:* Tricco AC, Lillie E, Zarin W, O'Brien KK, Colquhoun H, Levac D, et al. PRISMA Extension for Scoping Reviews (PRISMAScR): Checklist and Explanation. Ann Intern Med. 2018;169:467–473. doi: 10.7326/M18-0850.
